# Robust thalamic nuclei segmentation from T1-weighted MRI using polynomial intensity transformation

**DOI:** 10.1101/2024.01.30.24301606

**Authors:** Julie P. Vidal, Lola Danet, Patrice Péran, Jérémie Pariente, Meritxell Bach Cuadra, Natalie M. Zahr, Emmanuel J. Barbeau, Manojkumar Saranathan

**Affiliations:** CNRS, CerCo (Brain and Cognition Research Center) - Paul Sabatier University, Toulouse, France; INSERM, ToNiC (Toulouse NeuroImaging Center) – Paul Sabatier University, Toulouse, France; Purpan Hospital, Toulouse University Hospital Center, Neurology Department, Toulouse, France; CIBM (Center for Biomedical Imaging) – Radiology Department, Lausanne University and University Hospital, Lausanne, Switzerland; Department of Psychiatry & Behavioral Sciences - Stanford University School of Medicine, Stanford, California, USA; Department of Radiology - University of Massachusetts Chan Medical School, Worcester, Massachusetts, USA

**Keywords:** Thalamus, thalamic nuclei segmentation, THOMAS, structural imaging

## Abstract

Accurate segmentation of thalamic nuclei, crucial for understanding their role in healthy cognition and in pathologies, is challenging to achieve on standard T1-weighted (T1w) magnetic resonance imaging (MRI) due to poor image contrast. White-matter-nulled (WMn) MRI sequences improve intrathalamic contrast but are not part of clinical protocols or extant databases. In this study, we introduce histogram-based polynomial synthesis (HIPS), a fast preprocessing transform step that synthesizes WMn-like image contrast from standard T1w MRI using a polynomial approximation for intensity transformation. HIPS was incorporated into THalamus Optimized Multi-Atlas Segmentation (THOMAS) pipeline, a method developed and optimized for WMn MRI. HIPS-THOMAS was compared to a convolutional neural network (CNN)-based segmentation method and THOMAS modified for T1w images (T1w-THOMAS). The robustness and accuracy of the three methods were tested across different image contrasts (MPRAGE, SPGR, and MP2RAGE), scanner manufacturers (PHILIPS, GE, and Siemens), and field strengths (3T and 7T). HIPS-transformed images improved intra-thalamic contrast and thalamic boundaries, and HIPS-THOMAS yielded significantly higher mean Dice coefficients and reduced volume errors compared to both the CNN method and T1w-THOMAS. Finally, all three methods were compared using the frequently travelling human phantom MRI dataset for inter- and intra-scanner variability, with HIPS displaying the least inter-scanner variability and performing comparably with T1w-THOMAS for intra-scanner variability. In conclusion, our findings highlight the efficacy and robustness of HIPS in enhancing thalamic nuclei segmentation from standard T1w MRI.

## 1. Introduction

The thalamus is a deep brain structure on either side of the third ventricle, comprised of multiple nuclei (Morel et al., 1997). Physiologically, these nuclei are often classified as first order nuclei (e.g. lateral and medial geniculate nuclei), relaying sensory or motor information to the cortex, higher order nuclei (e.g. pulvinar) involved in cognition through cortico-thalamo-cortical circuits (Sherman, 2007) and the thalamic reticular nucleus. Several neurodegenerative, neurological, and neuropsychiatric conditions involve the thalamic nuclei such as alcohol use disorder (Zahr et al., 2020), schizophrenia (Andreasen, 1997), Alzheimer’s disease (Braak & Braak., 1991), chronic pain syndrome (Gustin et al., 2011), epilepsy (Fisher et al., 2010) and stroke (Danet et al., 2015). Thus, visualization and characterization of thalamic nuclei are crucial in understanding their function as well as their relationship in the healthy brain or in pathological conditions. This need is even more critical for pathologies where clinical care is based on accurate targeting of specific nuclei, such as deep brain stimulation (DBS) for the treatment of essential tremor (Koller et al., 2001), chronic pain syndrome (Owen et al., 2006) or drug-resistant epilepsy (Fisher et al., 2010). Other therapeutic approaches targeting specific thalamic nuclei include Magnetic Resonance Guided Focused Ultrasound thalamotomy (MRgFUS), gamma knife surgery, radio frequency surgery, and microsurgical resection (Benabid et al., 1997; Cinalli et al., 2018).

Segmentation of thalamic nuclei is challenging due to their poor contrast on standard T1 and T2 weighted (T1w, T2w) MRI, the most commonly used pulse sequences in routine neuroimaging protocols. To partly address this issue, atlases such as the Schaltenbrand-Warren and the Morel atlas (Morel et al., 1997; Schaltenbrand & Warren, 1977) have been employed. These atlases have been used for manual delineations of nuclei on MRI data as well as in neurosurgical targeting using stereotactic coordinates (Sanborn et al., 2009). Manual delineation of thalamic nuclei requires special expertise, is time consuming, and is therefore not ideal for analysis of large datasets. A three-dimensional reconstruction of the Morel Atlas in MNI space has been developed to help automate the segmentation of thalamic nuclei (Jakab et al., 2012). However, such atlas-based approaches ignore important inter-individual and inter-thalamic variability of the size, shape, and location of thalamic nuclei, leading to compromised accuracy.

To address the issue of automated thalamic nuclei parcellation at a subject-level and exploit image contrast, several approaches have been explored. One of the earliest methods, proposed by Behrens et al. (2003), used probabilistic tractography from Diffusion Tensor Imaging (DTI) data, mapping structural connectivity between different thalamic regions and specific cortical regions based on white matter anisotropy (e.g. mediodorsal nucleus to prefrontal cortex, lateral geniculate nucleus to visual cortex and so on), resulting in 7 thalamic regions corresponding to the 7 seed regions. Other diffusion MRI based methods have used local information from the diffusion tensor at a voxel level to parcellate the thalamus. Mang et al. (2012) used k-means clustering of the dominant diffusion orientation while Battistella et al. (2017) used k-means clustering of the spherical harmonic coefficients of the orientation distribution function to parcellate the thalamus into 6 regions. Since the thalamus is mainly composed of isotropic grey matter, the direction information computed from the diffusion tensor tends to be noisy. Further, the spatial resolution limitations of the underlying echo planar imaging acquisition results in a small number of clusters rather than precise, anatomically defined nuclei. Functional MRI based segmentation approaches have also been proposed using the idea of functional connectivity to cortical ROIs employing seed-based (Zhang et al., 2008), or Independent Components Analysis (ICA) (Hale et al., 2015). These also resulted in 6 thalamic regions corresponding to structural connectivity results of Behrens et al. In contrast, to parcellate the thalamus into 15 clusters, Kumar et al. (2017) proposed an ICA based functional parcellation while Van Oort et al. (2018) used time courses of instantaneous connectivity. Other methods based on susceptibility weighted imaging or quantitative susceptibility mapping (Liu et al., 2020; Deoni et al., 2005) have typically relied upon manual segmentation and have been limited to targeting the VIM nucleus for DBS treatment of movement disorders.

Images from structural (anatomic) MRI methods such as T1w Magnetization Prepared Rapid Gradient Echo (MPRAGE) have high spatial resolution (typically 1mm isotropic), minimal distortion, and are commonly used for cortical segmentation but rarely used for thalamic nuclei segmentation due to poor intra-thalamic nuclear contrast. Iglesias et al. developed a probabilistic atlas that combined manual delineations from in vivo T1w MPRAGE data and ex-vivo histological data (Iglesias et al., 2018). The Bayesian segmentation algorithm that uses this atlas to segment T1w MRI is part of the Freesurfer package. Variants of MPRAGE such as white-matter-nulled (WMn) (Tourdias et al., 2014), and grey-matter-nulled (GMn) (Magnotta et al., 2000) MPRAGE imaging significantly improves intra-thalamic contrast and permit better delineation of thalamic nuclei. A multi-atlas method called THalamus Optimized Multi-Atlas Segmentation (THOMAS) (Su et al., 2019) has been proposed that uses 20 WMn-MPRAGE prior datasets acquired at 7T and segmented manually using the Morel atlas as a guide combined with a joint-label fusion algorithm for thalamic nuclei parcellation of WMn-MPRAGE data. This method (which we call WMn-THOMAS) divided the thalamus into 11 nuclei per hemisphere and was validated against manual segmentation.

While WMn-THOMAS has been used in several studies examining the role of thalamic nuclei in alcohol use disorder and multiple sclerosis (Zahr et al., 2020; Su et al., 2020), WMn-MPRAGE sequences are neither part of commonly used clinical protocols nor available in existing data repositories like ADNI or OASIS. To segment conventional MPRAGE T1w data, THOMAS was recently modified to use mutual information (MI) instead of cross-correlation (CC) as the nonlinear registration metric (Bernstein et al., 2021; Pfefferbaum et al., 2023) and a majority voting algorithm for label fusion (which we call T1w-THOMAS). While this method achieved good accuracy compared to WMn-MPRAGE for larger nuclei such as the mediodorsal or pulvinar, it was less accurate for segmentation of the smaller centromedian and habenular nuclei. One reason for suboptimal performance of the modified T1w-THOMAS method could be due to poor intrathalamic contrast and thalamic boundaries on standard T1w contrast images. To address this limitation, a novel deep learning-based method (Umapathy et al., 2021) was proposed, that first synthesized WMn-MPRAGE-like images from T1w data and then segmented the synthetic WMn-like data, using two separately trained convolutional neural networks (CNN). This CNN-based synthesis approach was shown to improve accuracy compared to direct segmentation of T1w data (using a different CNN trained directly on 3T T1w data). However, the adoption of this method has been limited due to the necessity of training the CNNs on new types of data (e.g. different field strengths or scanner manufacturer). Furthermore, this CNN training process is time consuming and often not possible due to lack of T1w and WMn-MPRAGE data acquired concurrently on the same subjects.

Inspired by the promising CNN-based WMn synthesis approach of Umapathy et al. (2021) and leveraging the contrast benefits of WMn-THOMAS, we introduce here a new pre-processing transform step, Histogram-based Polynomial Synthesis (HIPS), that enables robust and improved thalamic nuclei segmentation by synthesizing WMn-like images from T1w MRI using a simple polynomial function for intensity transformation. To test the performance of HIPS-THOMAS against T1w-THOMAS and CNN-based segmentation, quantitative performance metrics including Dice, volume errors, and inter-scanner/intra-scanner variability were used to assess performance across differing contrast (MPRAGE, SPGR, MP2RAGE), scanner manufacturers (Philips, GE, Siemens), and field strengths (1.5T, 3T, 7T).

## 2. Methods

### 2.1 Histogram-based polynomial synthesis (HIPS): motivation

The main motivation of HIPS is to enable accurate thalamic nuclei segmentation from T1w images. As WMn-MPRAGE images were demonstrated to possess a higher intrathalamic contrast (Su et al., 2019) and generate more accurate segmentation (Umapathy et al. 2021) compared to T1w images, the goal was to generate WMn-like images from T1w data prior to THOMAS segmentation. Examining the two image contrasts revealed that the two images are nearly the “reverse” of each other with cerebrospinal fluid appearing dark in one and bright in another while the white matter appears bright in one and dark in the other. This suggested an intuitively simple “contrast reversal” scheme, which was implemented as a *1-x* operation, with x corresponding to normalized image intensity values. The normalization procedure ensured that images from different subjects or scanners could be transformed in a similar fashion, independent of scaling factors and overall intensity differences and is described in detail in the next section. This *1-x* linear scheme was applied to T1w images to generate WMn-like images. However, plots of normalized T1w vs WMn image intensities revealed that a linear fit was suboptimal and further corroborated by the “darkened” appearance on the contrast reversed image (first column of Supplementary Figure 1). As a result, a polynomial function was used to better fit the nonlinear data as described below.

### 2.2 HIPS methodology

The main steps of HIPS preprocessing are as follows:

a. Image preparation: Images were first cropped to focus on the bilateral thalami and avoid the skull or fat/skin tissue. Note that this cropping was done automatically using a cropped mask template warped from template space as described in Su et al. (2019) and is already part of the THOMAS pipeline.
b. Image normalization: Image histograms were computed using a representative central axial slice of the cropped dataset, encompassing both thalami. To render the contrast transformation step independent of scanner type and subjects, T1-MPRAGE images were normalized using the WM signal and WMn-MPRAGE images were normalized using the CSF signal, both extracted from their respective image histograms, prior to fitting. To perform the normalization, the mode of the tissue of interest (WM in T1w, CSF in WMn-MPRAGE) was first computed from the histogram and the highest value shared by at least 1% of voxels exceeding the mode was used for the normalization (voxel value/normalizing value). This approach is like the WhiteStripe intensity normalization method (Shinohara et al., 2014) and is effective in removing the high intensity tail corresponding to artifacts and outlier intensities (Sun et al., 2015). Note that conventional contrast scaling based on maximum and minimum image intensity did not work due to noise and the presence of tissue other than WM/GM/CSF which caused variability in contrast (and hence, image appearance) across subjects or scanners.
c. Polynomial transformation: The desired contrast transformation is essentially estimating a function that optimally maps the normalized T1w space to normalized WMn space. The general form of the function was 1 + ax + bx^2^ + cx^3^ + dx^4^ with d = 0 for 3^rd^ order, c and d = 0 for 2^nd^ order, and a=-1, b, c, and d = 0 for linear. See Section 2.3 for details on determining optimal order and coefficients.
d. Contrast stretching and rescaling: To further maximize the image contrast, a contrast stretching step which rescales intensities within the 2 and 98 percentile range of the input image values was performed. Finally, images are rescaled to the highest WM value (mode) computed in the Image normalization step to restore the image intensity ranges (corresponding to the original images prior to normalization).

### 2.3 HIPS: parameter optimization and validation

The optimal polynomial order and coefficients for HIPS were estimated using a training set of 10 T1-MPRAGE and WMn-MPRAGE datasets acquired concurrently from the same subjects on a Philips 3T scanner. Following the cropping step to focus on the bilateral thalami, data from the two image contrasts were affinely registered using Advanced Normalization Tools (ANTS) (Avant, Tustison & Song, 2009) with WMn-MPRAGE as fixed image and T1-MPRAGE as moving image. Polynomial functions of orders 1 (i.e. linear) to 4 (quartic) were tested using a curve fitting function (*scipy.optimize.curve*_*fit*, python 3.9). For each order, a single aggregate function derived from the 10 training subjects was applied to a test set of normalized T1w images (10 Philips 3T, 9 GE 3T, 10 Siemens 3T, and 8 Siemens 7T datasets) to generate the corresponding synthetic WMn images. These WMn-like images were first visually evaluated against the corresponding native WMn-MPRAGE data using a kernel density estimation (KDE) plot (seaborn.kdeplot, python 3.9). The density plot approximates the underlying probability density function that generated the data but rather than using discrete bins like a histogram, it smooths the observations with a Gaussian kernel, producing a continuous density estimate for better visualization. A thin diagonal line on a density plot indicates an almost perfect concordance between the two normalized intensities of images compared. The optimality of the transform functions was also assessed using two quantitative metrics-Structural Similarity Index (SSI) and Mean Square Error (MSE) and the best function was used for all subsequent processing. Supplementary Figure 1 shows the effects of contrast transformation on an example case using the 4 orders.

### 2.4 The HIPS-THOMAS segmentation pipeline

HIPS-THOMAS is a variant of the THalamus Optimized Multi Atlas Segmentation (THOMAS) method (Su et al., 2019) and is shown in Figure 1. The input T1w image is first cropped to cover both thalami which removes outliers from the skull/subcutaneous fat. Following the cropping step, HIPS preprocessing is applied as described in detail in Section 2.2 comprising of image normalization, application of the optimized polynomial contrast transformation, and finally contrast-stretching and rescaling. This results in a WMn-like version of the cropped input T1w image. This HIPS transformed cropped image is then nonlinearly registered (”Warp R”) to a cropped average brain template from 20 WMn-MPRAGE priors using the cross correlation metric (”CC” in blue, Fig. 1). This nonlinear warp is inverted (“R-^1^”, Fig. 1) and combined with the 20 precomputed warps from priors to the template (“W_piT_”, Fig. 1) to put the 20 manual segmentation labels in the input space. These 20 labels were then combined using a joint label fusion algorithm to generate the output’s final parcellation. The segmented thalamic nuclei include Anteroventral (AV), Ventral anterior (VA), Ventral lateral anterior (VLa), Ventral lateral posterior (VLp), Ventral posterolateral (VPL), Pulvinar (Pul), Lateral geniculate (LGN), Medial geniculate (MGN), Centromedian (CM), Mediodorsal-Parafascicular (MD-Pf) in addition to the Habenula (Hb) and the Mammillothalamic tract (MTT) and are shown in Table 1.

**Fig. 1:**
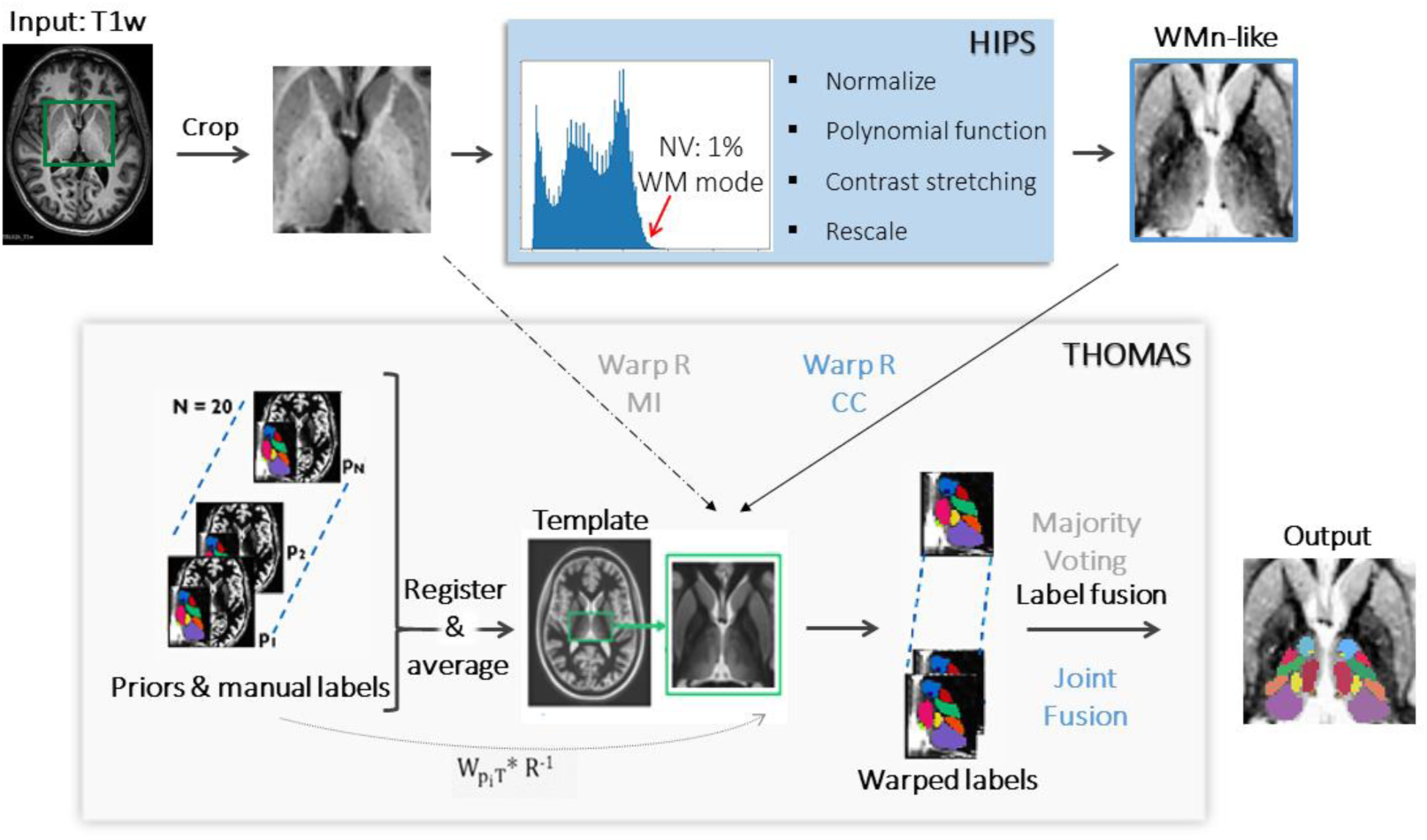
The proposed HIPS-THOMAS pipeline. HIPS pre-processing includes normalization of the cropped T1w input, application of a polynomial function to generate a WMn-like image, and a contrast stretching and a rescaling step. The WMn-like cropped input is fed into the THOMAS pipeline as opposed to the original cropped T1w image. Note that for HIPS-THOMAS, the nonlinear warp R uses a cross-correlation metric (CC in blue) and the label fusion step uses joint label fusion (in blue) as opposed to mutual information metric (MI in grey) and majority voting (in grey). NV: normalizing value.

**Table 1:**
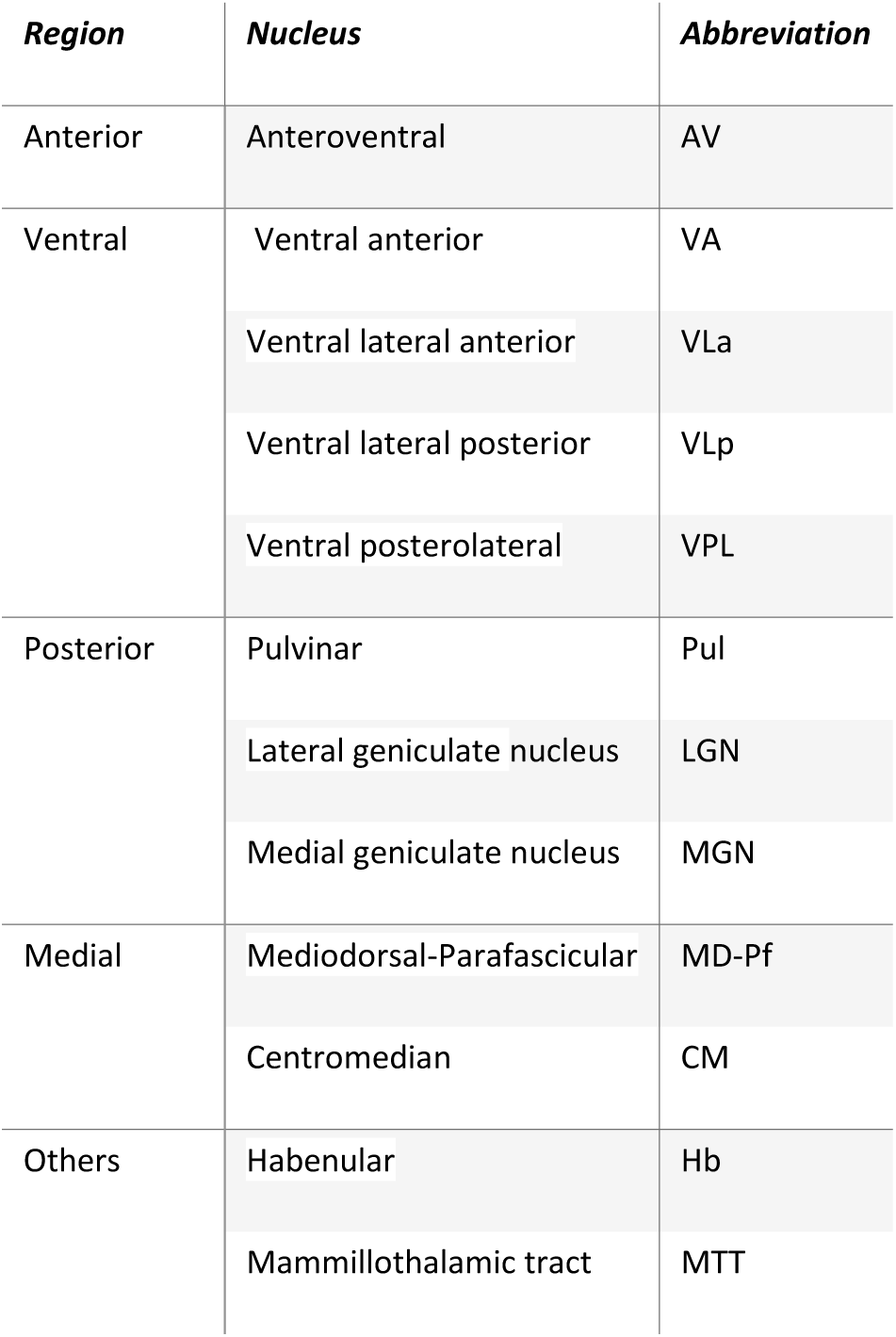
List of thalamic nuclei segmented by THOMAS following the nomenclature of Morel.

The two notable differences between HIPS-THOMAS and T1w-THOMAS are the use of cross-correlation metric (”CC” in blue, Fig. 1) for ANTs nonlinear registration in calculating the Warp R instead of mutual information metric (“MI” in grey, Fig. 1) and the use of joint label fusion instead of majority voting in the final label fusion step.

### 2.5 Convolutional Neural Network (CNN) based segmentation

In addition to T1w-THOMAS and HIPS-THOMAS, the dual CNN method of Umapathy et al. (2021) was also used for segmentation of T1w data. This approach uses two cascaded 3D CNNs-WMn-MPRAGE-like images are first synthesized from T1w images using a contrast synthesis CNN and these images are then processed using another CNN to yield thalamic nuclei parcellations. The synthesis network was trained using patches from registered pairs of T1w and WMn-MPRAGE images acquired from the same subjects on 3T GE and Siemens scanners as described in Umapathy et al (2021); the segmentation network was trained using WMn-THOMAS data.

### 2.6 Datasets and evaluation metrics

The datasets used in the analysis comprised of 12 subjects acquired on a Siemens 3T scanner with T1w MPRAGE, 19 subjects acquired on a GE 3T scanner with 3D SPGR, and 18 subjects acquired on a Philips 3T scanner with T1w MPRAGE. WMn-MPRAGE acquired on each of these subjects were also available for comparisons. In addition, 8 datasets acquired on a Siemens 7T scanner using Magnetization-Prepared 2 Rapid Acquisition Gradient Echo (MP2RAGE) sequence were also analyzed. All experimental protocols including data acquisition were approved by institutional review board guidelines (University of Arizona for Siemens 3T data; Stanford and SRI International for GE 3T data; Comité de protection des personnes Ile-de-France IV for Philips 3T data; Commission cantonale d’éthique de la recherche sur l’être humain (CER-VD) for Siemens 7T data) and all data were acquired after obtaining prior written informed consent from the participants in accordance with the Declaration of Helsinki. Additional information about each sequence can be found in Supplementary Table 1. Segmentation performance of T1w-THOMAS, HIPS-THOMAS, and CNN were compared for different-

i. T1w contrast (MPRAGE, SPGR, MP2RAGE)
ii. scanner manufacturers (Siemens, GE, Philips)
iii. field strengths (3T, 7T).

In the absence of “gold standard” manual segmentations, segmentations were compared to the “silver standard” WMn-THOMAS, using Dice coefficients which is commonly used to assess overlap of segmentations: 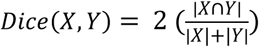 where |X| and |Y| are the cardinality of ground truth X and segmentation Y i.e. |X| + |Y| is the total number of voxels in each image X and Y while |X ∩ Y| is the cardinality of the intersection between ground truth X and segmentation Y i.e. the number of voxels that are present in both nuclei segmentations X and Y. Percentage volume error is also computed as 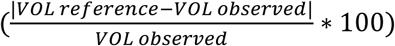 for each nucleus. Statistical significance was determined using t-tests with a Bonferroni correction (p-value / total number of comparisons = 0.05/13) to correct the p-values for multiple comparisons.

### 2.7 Analysis of the Frequently Traveling Human Phantom (FTHP) MRI dataset

To assess the inter-scanner and intra-scanner variability of the different methods, we used a subset of the FTHP MRI dataset (Opfer et al., 2023). Briefly, this dataset comprises of T1w MRI from a single healthy male volunteer (age around 50 years old) scanned on 116 different MRI scanners. We used 3 scanner manufacturers (Philips, Siemens, and GE), 2 field strengths (1.5T and 3T) and 4 different sites/scanner models for each manufacturer with 3 repeat scans at each site, resulting in a total of 72 scans. Intra-scanner variability was assessed from volume residuals by subtracting the mean volume of the 3 repeat scans acquired from the same scanner from individual volumes as in Opfer et al. (2023)., resulting in a vector of length 72. Inter-scanner variability was similarly assessed by first averaging the volumes from the three repeat scans and then computing residuals on the resulting vector of length 24.

## 3. Results

### 3.1 HIPS parameter optimization

For the four polynomial orders considered, 2^nd^ and 3^rd^ order functions performed the best, based on SSI and MSE metrics. The only exception was Siemens 7T where the linear function performed the best. These results are summarized in Supplementary Table 2. The 3rd order polynomial equation was selected for use as we hypothesized more accuracy by considering the three tissues of interest (white matter, gray matter, cerebrospinal fluid). A plot of T1w vs. WMn-MPRAGE normalized image intensities (blue dots) for a representative Philips 3T subject is shown in Figure 2a with the best fit 3^rd^ order curve shown in green. Individual best fit 3^rd^ order curves for 10 Philips 3T subjects are shown in Figure 2b with the single curve aggregated from all 10 subjects shown overlaid in red. The optimal curves for each order are shown below:

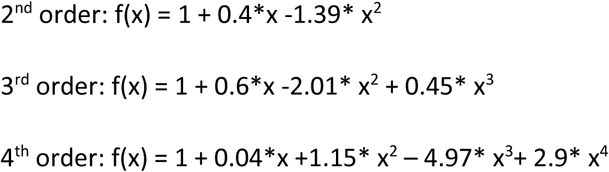

**Fig. 2:**
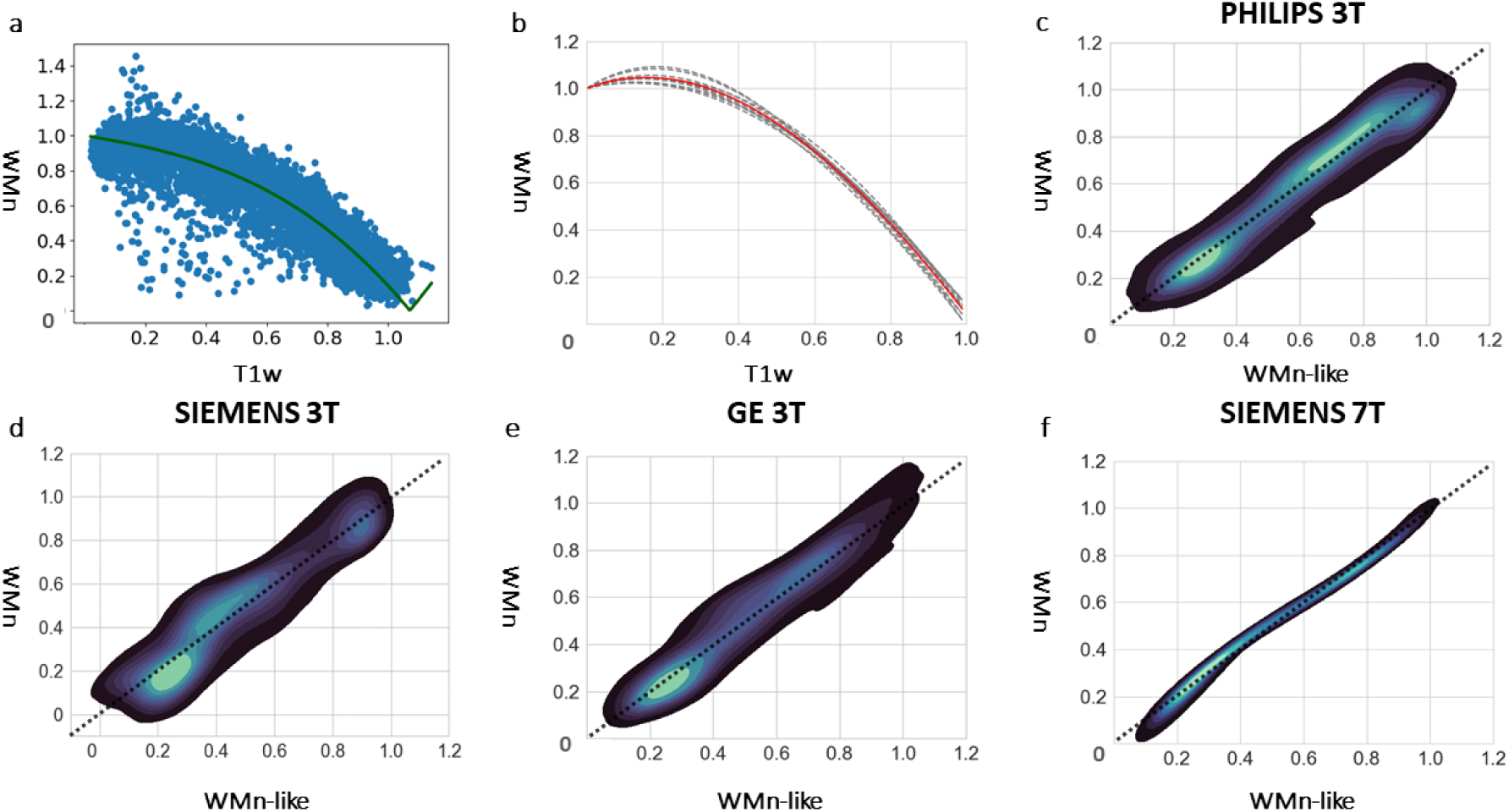
HIPS formulation and performance. (a) Normalized intensity plot between WMn-MPRAGE and T1-MPRAGE data from a Philips 3T subject (blue dots) and a 3^rd^ order polynomial fit (green line). (b) The 10 curves resulting from the individual fitting on 10 Philips data cases (dashed gray) and the resulting aggregated function (red line in a, b). Density plots between normalized WMn-MPRAGE and synthesized WMn-MPRAGE data using the same aggregated function on an example subject from Philips 3T (c), Siemens 3T (d), GE 3T (e), and Siemens 7T (f). The black dashed unity line represents perfect concordance between images. Both axes show normalized voxel intensities.

Since this function was derived from normalized data, it was applicable across all subjects from different scanners and field strengths. Density plots between native cropped central slice WMn-MPRAGE and WMn-like synthesized images obtained through the application of the aggregated function on an example case from each of the 4 scanner types are shown in Figures 2c-f. The linearity of the density plots attests to the quality and robustness of the synthesis, with the Siemens 7T showing the most concordance (minimal dispersion from the unity line). To confirm the ability of this Philips data-based 3^rd^ order function to be optimal for all scanner-types, we compared this function to a function derived from mixed data (10 Philips 3T MPRAGE, 9 GE 3T SPGR, 10 Siemens 3T MPRAGE and 8 Siemens 7T MP2RAGE cases) and found no significant differences in SSI or MSE (Supplementary Table 3). This is further confirmed by the similarity of transformations between resulting WMn-like images after the application of the Philips equation vs. mixed equation on T1w-MRI (Supplementary Figure 2).

### 3.2 Qualitative comparisons

Figure 3 shows acquired T1w and WMn-MPRAGE images as well as HIPS and CNN-synthesized WMn-like images for a Siemens 3T subject (a-d) and a GE 3T subject (e-h). The corresponding thalamic nuclei segmentations for the left side are also shown overlaid. Both the CNN and HIPS synthesized WMn images show improved intra-thalamic contrast (brighter signal in MD and pulvinar nuclei) and thalamic boundaries (white arrows) compared to T1w images. The CNN-synthesized images look less noisy with even better contrast compared to HIPS (d, h). T1w and WMn-MPRAGE images as well as HIPS and CNN-synthesized WMn-like images for a Philips 3T subject (i-l) and a Siemens 7T subject (m-p) are also shown in Figure 3. Note that the CNN was trained only using GE and Siemens 3T data as described earlier and the Philips and 7T represent a different scan manufacturer and field strength, respectively, as a test of robustness. HIPS-synthesized images look very similar to WMn-MPRAGE images and produce segmentations comparable to those of WMn-THOMAS for both Philips 3T and Siemens 7T subjects. The CNN method failed on more than a third of the Philips cases (Fig. 3l) and on all Siemens 7T cases (Fig. 3p) due to failures in the synthesis step, leading to poor performance of the subsequent segmentation CNN.

**Fig. 3:**
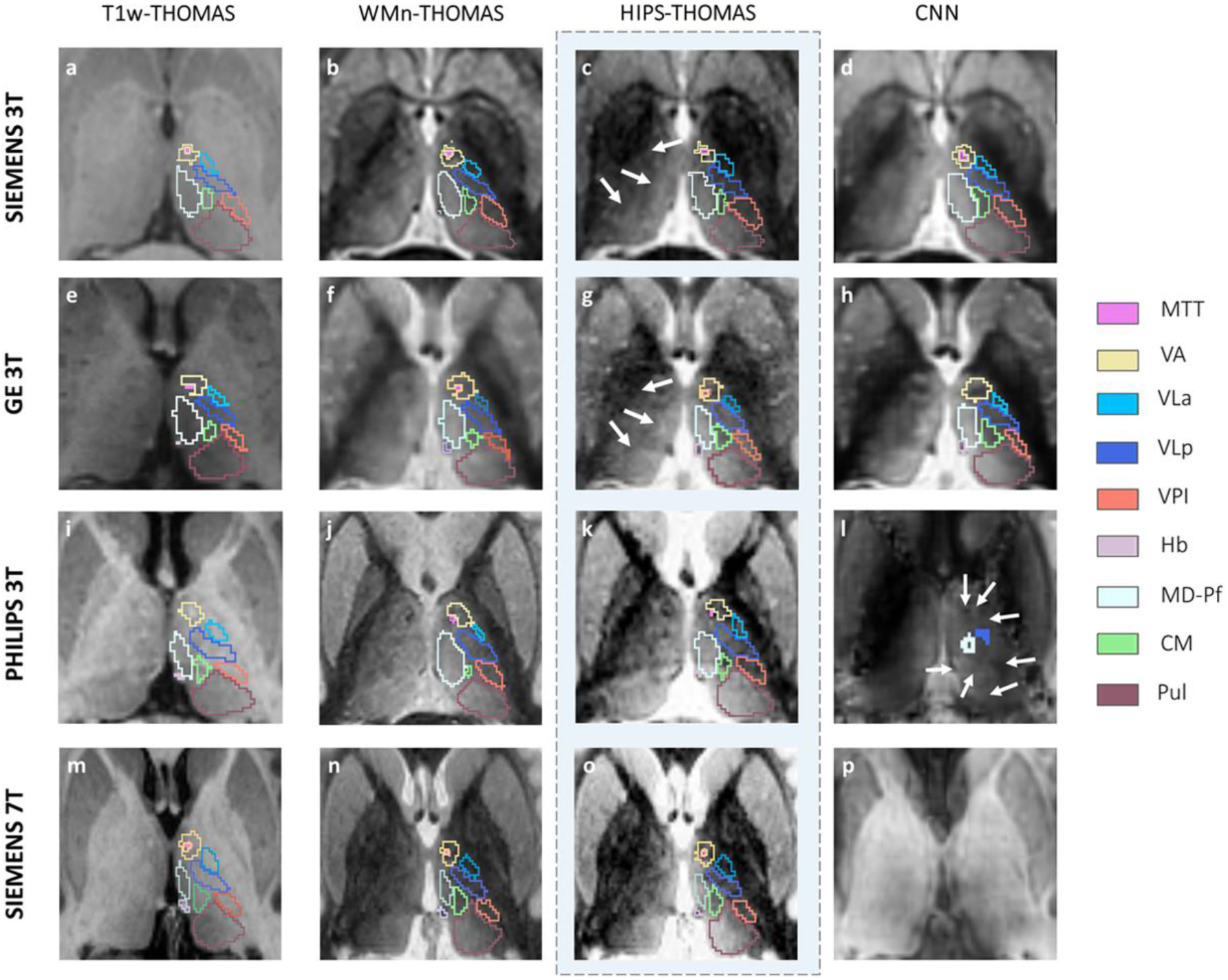
An axial slice from acquired T1w and WMn-MPRAGE as well as HIPS and CNN-synthesized WMn images for a Siemens 3T MPRAGE (a-d), GE 3T SPGR (e-h), Philips 3T MPRAGE (i-l) or Siemens 7T MP2RAGE (m-p) subject with the corresponding nuclei segmentations overlaid on the left thalamus. Note the improved intrathalamic contrast and thalamic boundaries (white arrows on c and g) in the synthesized WMn-like images produced by HIPS-THOMAS compared to the native T1w images. The failure of CNN synthesis can clearly be seen in panels l and p along with the failed or missing (white arrows in l) segmentations. Labels: Table 1

### 3.3 Quantitative assessments

For quantitative comparisons, mean Dice coefficients from Siemens 3T MPRAGE (n=12) and GE 3T SPGR (n=19) datasets were compared between T1w-THOMAS, HIPS-THOMAS, and CNN using THOMAS segmentation from WMn-MPRAGE as a silver-standard and the % improvement over T1w-THOMAS was computed. HIPS-THOMAS showed significantly improved Dice compared to T1w-THOMAS for 7/11 nuclei and the whole thalamus on both Siemens 3T and GE data (Figures 4-5). The raw data are tabulated in Supplementary Tables 4 and 5 for the Siemens 3T MPRAGE and the GE 3T SPGR datasets respectively. For HIPS-THOMAS, more than 10% increase in Dice for the VA, VLp, VPL, and LGN nuclei was observed on Siemens 3T MPRAGE and more than 15% increase in Dice for the VA, VLp, VPL, LGN, and CM nuclei was observed on GE 3T SPGR data. By contrast, the CNN performed better than T1w-THOMAS on 1/11 nuclei on Siemens 3T data and 6/11 on GE 3T data (“¤” in Fig. 4 and Fig. 5) with decreased Dice for several nuclei significant only for the MTT (Supp. Tab. 5). Notably, HIPS-THOMAS outperformed the CNN, with a higher mean and lower standard deviation, for many nuclei, especially the VLp and the MTT for both GE and Siemens data but also the VA, VPL, Pul, MGN, and MD on GE 3T data. Like the performance on GE and Siemens 3T, HIPS-THOMAS significantly improved Dice compared to T1w-THOMAS for 8/11 nuclei and the whole thalamus on Philips 3T data (Fig. 6) and 9/11 nuclei and the whole thalamus on Siemens 7T data (Fig. 7) with the corresponding data tabulated in Supplementary Table 6. More than 15% increase in Dice for the VA, VLp, MD nuclei was observed on Philips 3T data and more than 30% increase for the AV, VA, VLa, VLp, VPL, CM, MD nuclei on the Siemens 7T data. Only the Hb nucleus showed decreased Dice using HIPS-THOMAS compared to T1w-THOMAS on SIEMENS 7T (not statistically significant). Subjects who were scanned on 7T were also scanned on 3T and this data is reported in Supplementary Table 6. HIPS-THOMAS at 7T is substantially better (>15%) than at 3T on the VLa, LGN, and CM nuclei as well as the MTT.

**Fig. 4:**
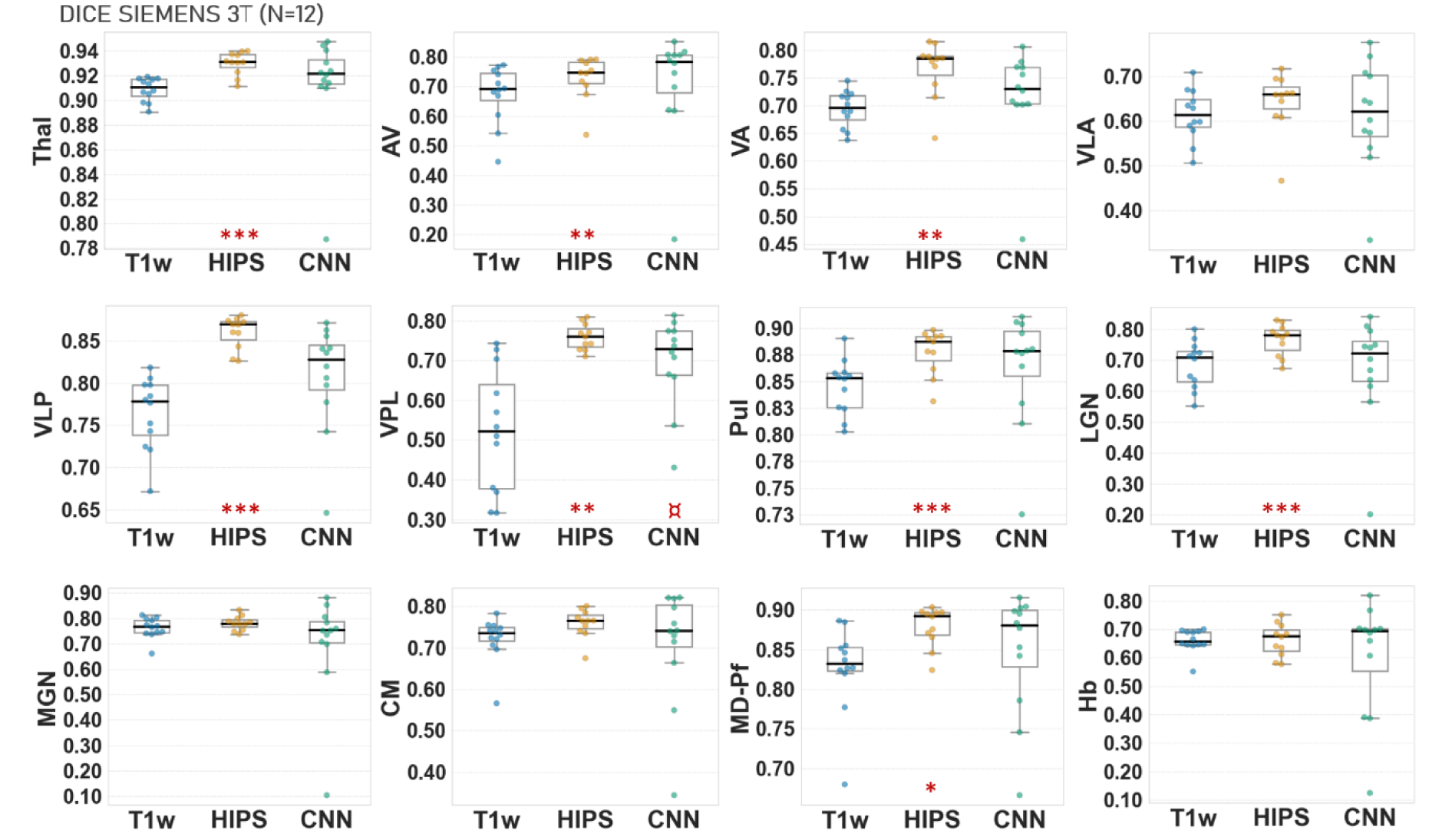
Box plots of T1w-THOMAS (T1w), HIPS-THOMAS (HIPS), and CNN segmentation’s Dice coefficients (compared against WMn-THOMAS segmentations) for 3T Siemens MPRAGE data (n=12). HIPS-THOMAS significantly improves Dice in whole thalamus and 7 nuclei compared to 1 nucleus for CNN. T-test between the Dice coefficients of the T1w-THOMAS vs. HIPS-THOMAS: P-values *<0.00385 (Bonferonni correction for multiple comparisons, 0.05/13) **<0.001 ***<0.0001. T1w-THOMAS vs. CNN: ¤ p<0.00385 (Bonferonni correction for multiple comparisons, 0.05/13), ¤¤ p<0.001, ¤¤¤ p<0.0001. Labels: cf. Table 1.

**Fig. 5:**
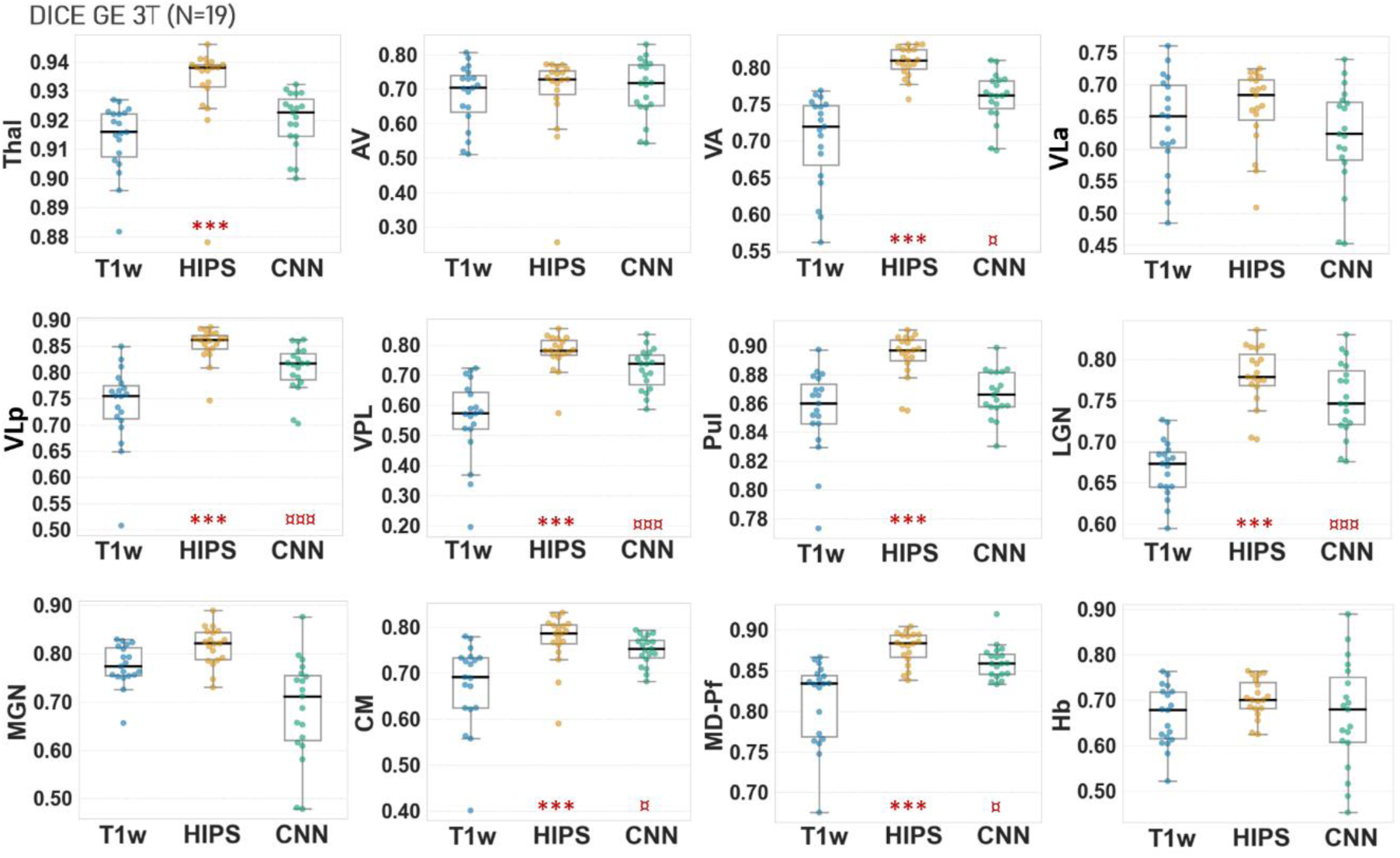
Box plots of T1w-THOMAS (T1w), HIPS-THOMAS (HIPS), and CNN segmentation’s Dice coefficients (compared against WMn-THOMAS segmentations) for GE 3T SPGR data (n=19). HIPS-THOMAS significantly improves Dice in whole thalamus and 7 nuclei compared to 6 nuclei for CNN. T-test between the Dice coefficients of the T1w-THOMAS vs. HIPS-THOMAS: P-values *<0.00385 (Bonferonni correction for multiple comparisons, 0.05/13) **<0.001 ***<0.0001. T1w-THOMAS vs. CNN: ¤ p<0.00385 (Bonferonni correction for multiple comparisons, 0.05/13), ¤¤ p<0.001, ¤¤¤ p<0.0001. Labels: cf. Table 1.

**Fig. 6:**
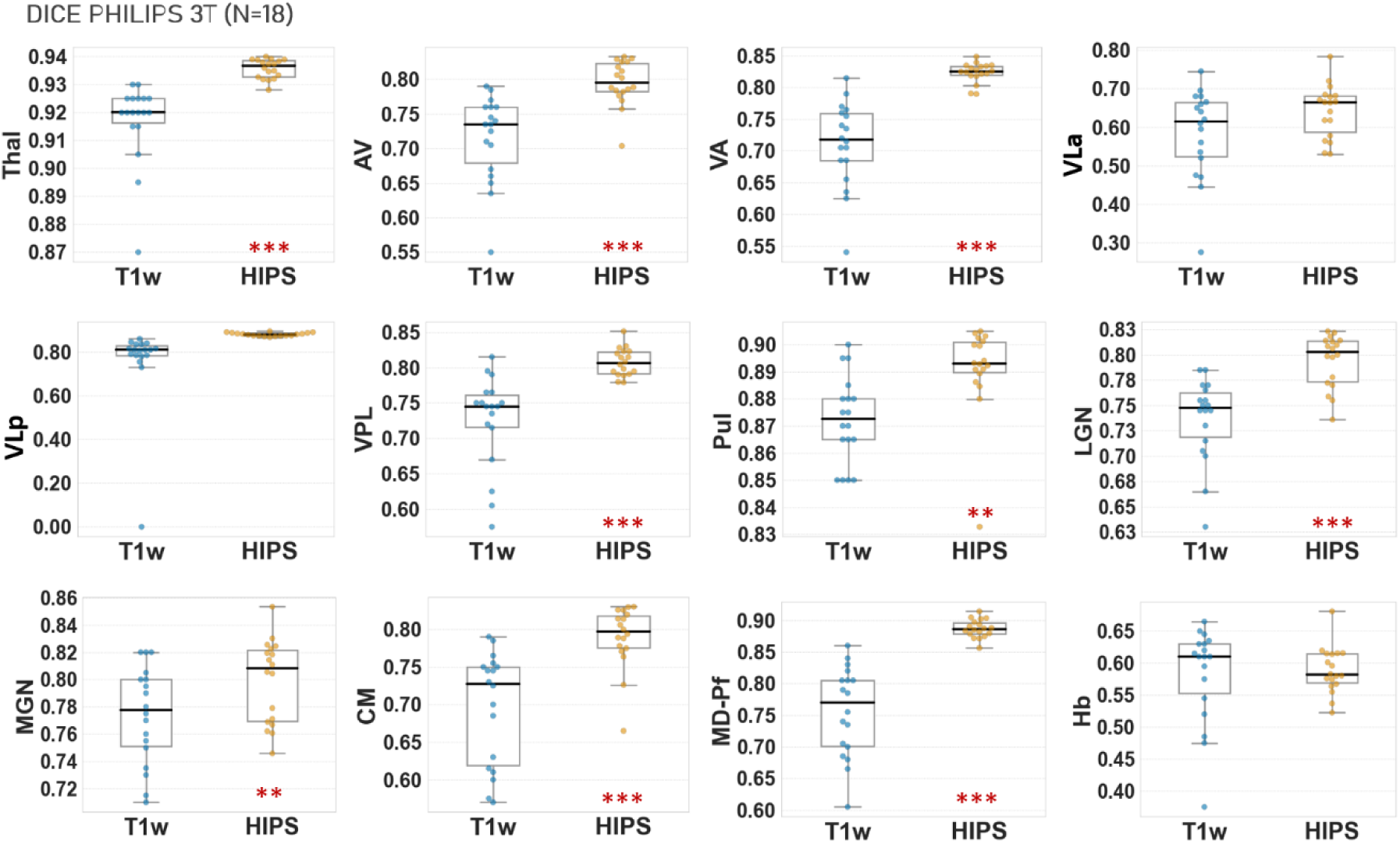
Box plots of T1w-THOMAS (T1w) and HIPS-THOMAS (HIPS) Dice coefficients (compared against WMn-THOMAS segmentations) for Philips 3T MPRAGE data (n=18). HIPS-THOMAS significantly improves Dice in whole thalamus and 8 nuclei. T-test between the DICE coefficients of the T1w-THOMAS VS HIPS-THOMAS: P-values *<0.00385 (Bonferonni correction for multiple comparisons, 0.05/13) **<0.001 ***<0.0001. Labels: cf. Table 1.

**Fig. 7:**
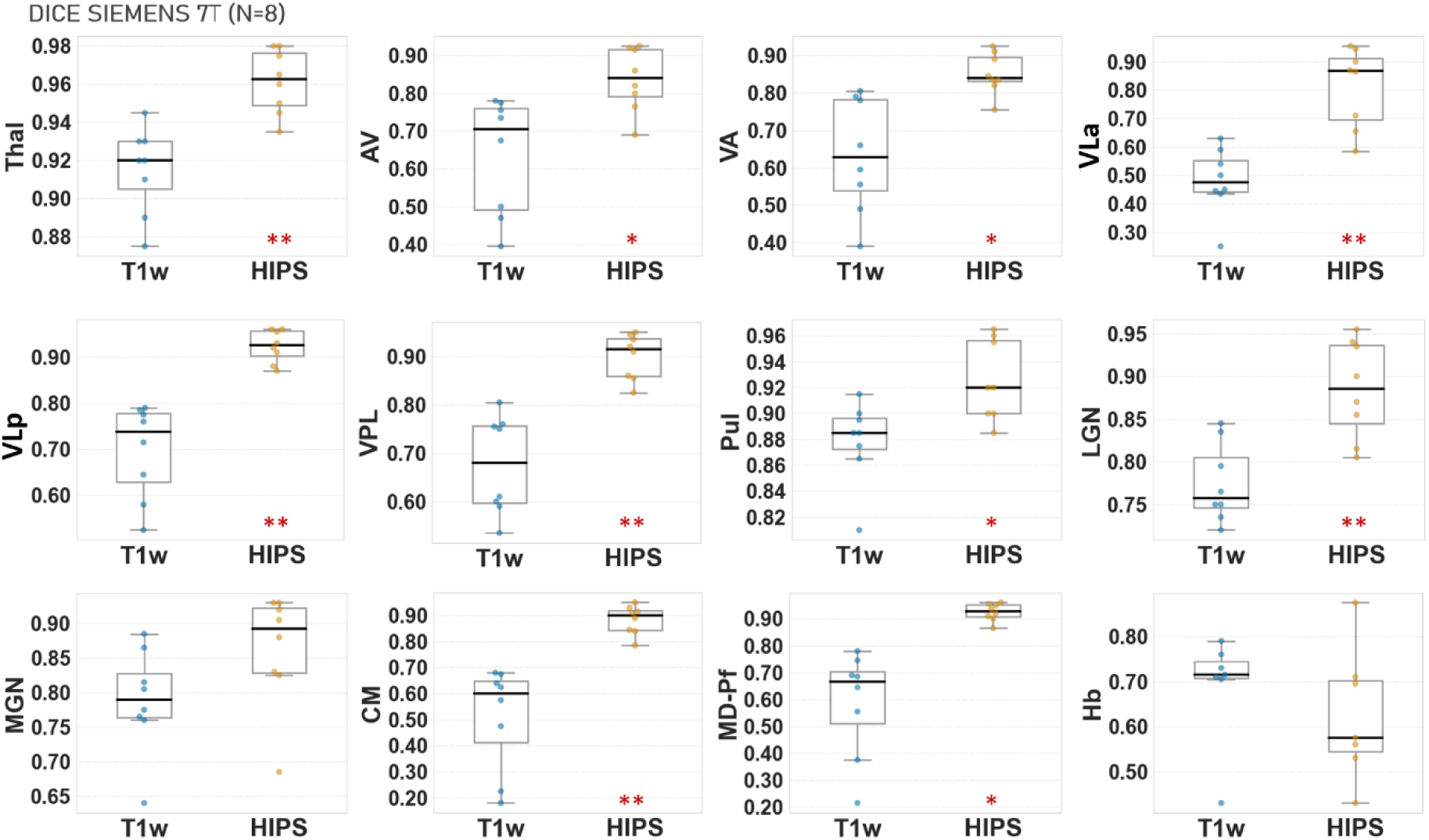
Box plots of T1w-THOMAS (T1w) and HIPS-THOMAS (HIPS) Dice coefficients (compared against WMn-THOMAS segmentations) for Siemens 7T MP2RAGE data (n=8). HIPS-THOMAS improves Dice in whole thalamus and 9 nuclei. T-test between the Dice coefficients of the T1w-THOMAS VS HIPS-THOMAS: P-values *<0.00385 (Bonferonni correction for multiple comparisons, 0.05/13) **<0.001 ***<0.0001. Labels: cf. Table 1.

Mean volume errors (expressed as percentage) of T1w-THOMAS, HIPS-THOMAS, and CNN segmentations compared to WMn-THOMAS segmentation are shown in Figure 8 for Siemens 3T (a), GE 3T (b), Philips 3T (c), and Siemens 7T (d) datasets. HIPS-THOMAS had the **lowest** error in 9 nuclei and the MTT and highest error in 2 nuclei for Siemens 3T data. A similar trend was also observed for the GE 3T data. T1w-THOMAS displayed the **highest** errors in 8 nuclei on Siemens 3T data and 7 nuclei on GE 3T data. CNN performance was either comparable to or slightly worse than HIPS except in the MTT and the AV nucleus on Siemens data, where it was substantially worse (higher error) and the AV nucleus on GE data, where it was substantially better (lower error). HIPS-THOMAS had lower mean errors on all nuclei for Philips 3T data and all except the Hb nucleus for Siemens 7T data.

**Fig. 8:**
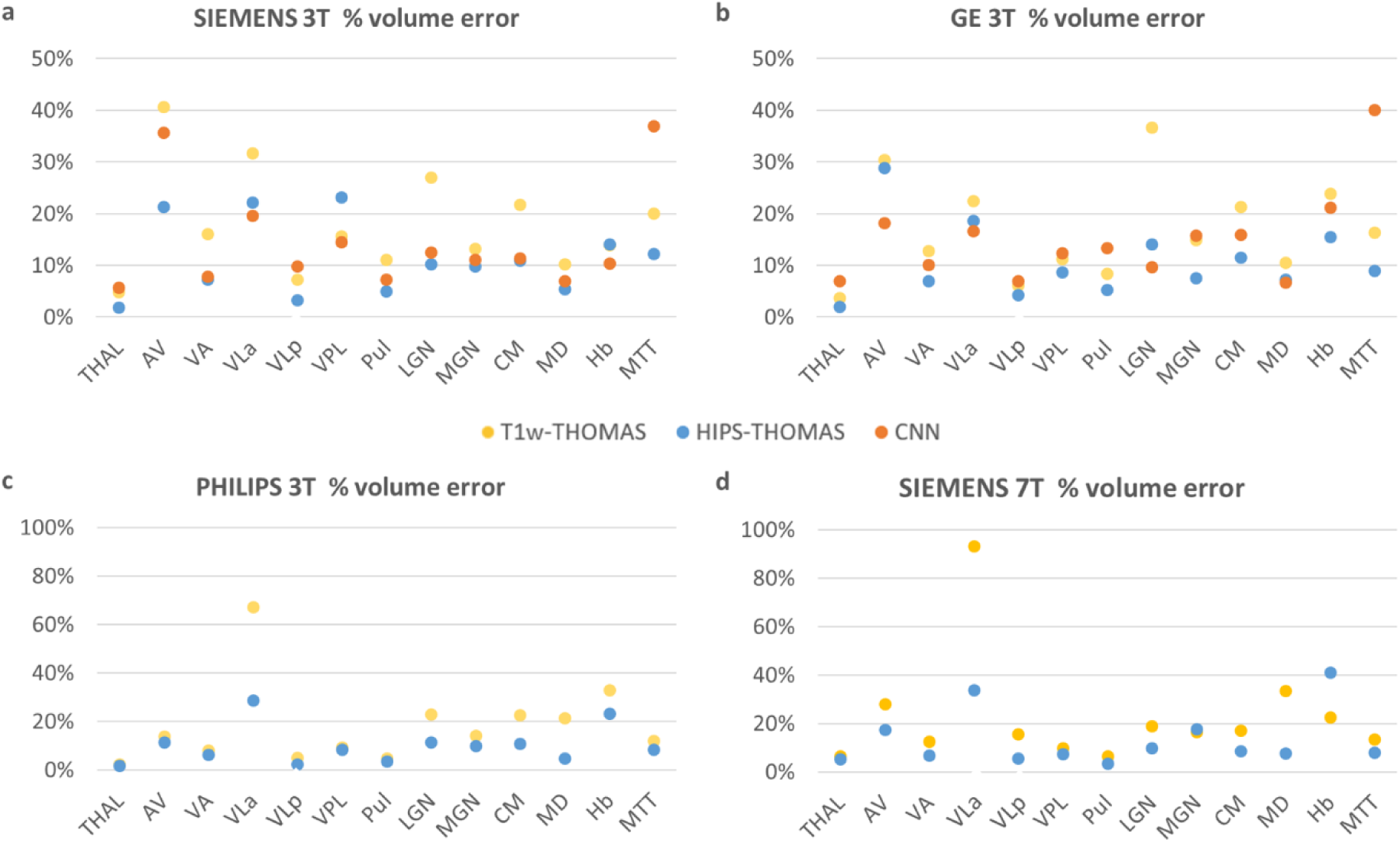
Mean volume error (%) of T1w-THOMAS, HIPS-THOMAS, and CNN segmentations, compared to WMn-THOMAS segmentations for Siemens 3T MPRAGE (a, n=12) and GE 3T SPGR (b, n=19) Philips 3T MPRAGE 3T (c, n=18) and Siemens 7T MP2RAGE (d, n=8) data. The general trend of HIPS < CNN < T1w THOMAS was observed for most nuclei at 3T. HIPS-THOMAS errors were lower than T1w-THOMAS for all nuclei but Hb for Siemens 7T data. Labels: cf. Table 1.

Figure 9 visually summarizes the improvements in Dice (%) and reduction in volume error % for HIPS-THOMAS compared to T1w-THOMAS for the 4 scanners (GE 3T, Siemens 3T, Philips 3T, and Siemens 7T). While 7T HIPS-THOMAS displayed the largest increase in Dice coefficients and reduction in volume errors, 3T showed at least 15% increase in Dice in ventral nuclei such as the VA and VPL across all scanners. Volume errors were also reduced for HIPS-THOMAS for all the scanners except for the VPL and Hb nuclei on GE 3T data and the Hb nucleus on Siemens 7T data (white regions).

**Fig. 9:**
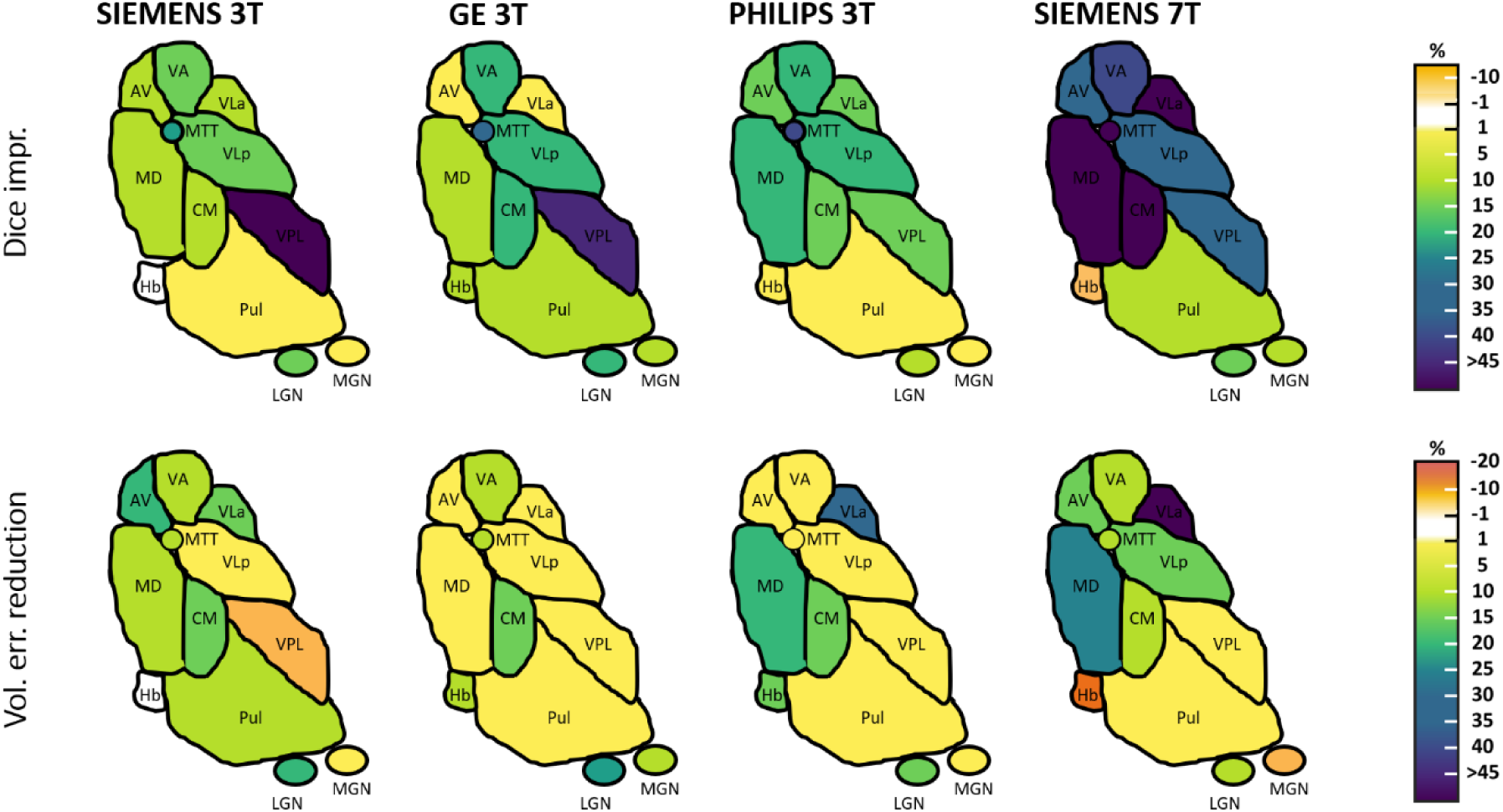
Graphical summary of improvement of mean Dice coefficients (%) (top row) and reduction of % volume error (bottom row) for each thalamic nucleus from Siemens 3T, GE 3T, PHILIPS 3T and Siemens 7T data using HIPS-THOMAS compared to T1w-THOMAS. Labels: cf. Method 2.2.

### 3.4 Inter- and intra-scanner variability

Figure 10 shows inter-scanner and intra-scanner variability for the whole thalamus (Thal), a small nucleus (AV) and a large nucleus (VLp). The results for all the thalamic nuclei are reported in Supplementary Table 7. The standard deviation of the residuals is reported on the top of each panel. HIPS-THOMAS had the least inter-scanner variability for the whole thalamus and 8/11 nuclei. It also had the least intra-scanner variability for the whole thalamus and 4/11 nuclei. In contrast, T1w-THOMAS had the least inter-scanner variability for 4/11 nuclei and least intra-scanner variability for 8/11 nuclei. CNN had the worst inter- and intra-scanner variability for the whole thalamus and all nuclei.

**Fig. 10:**
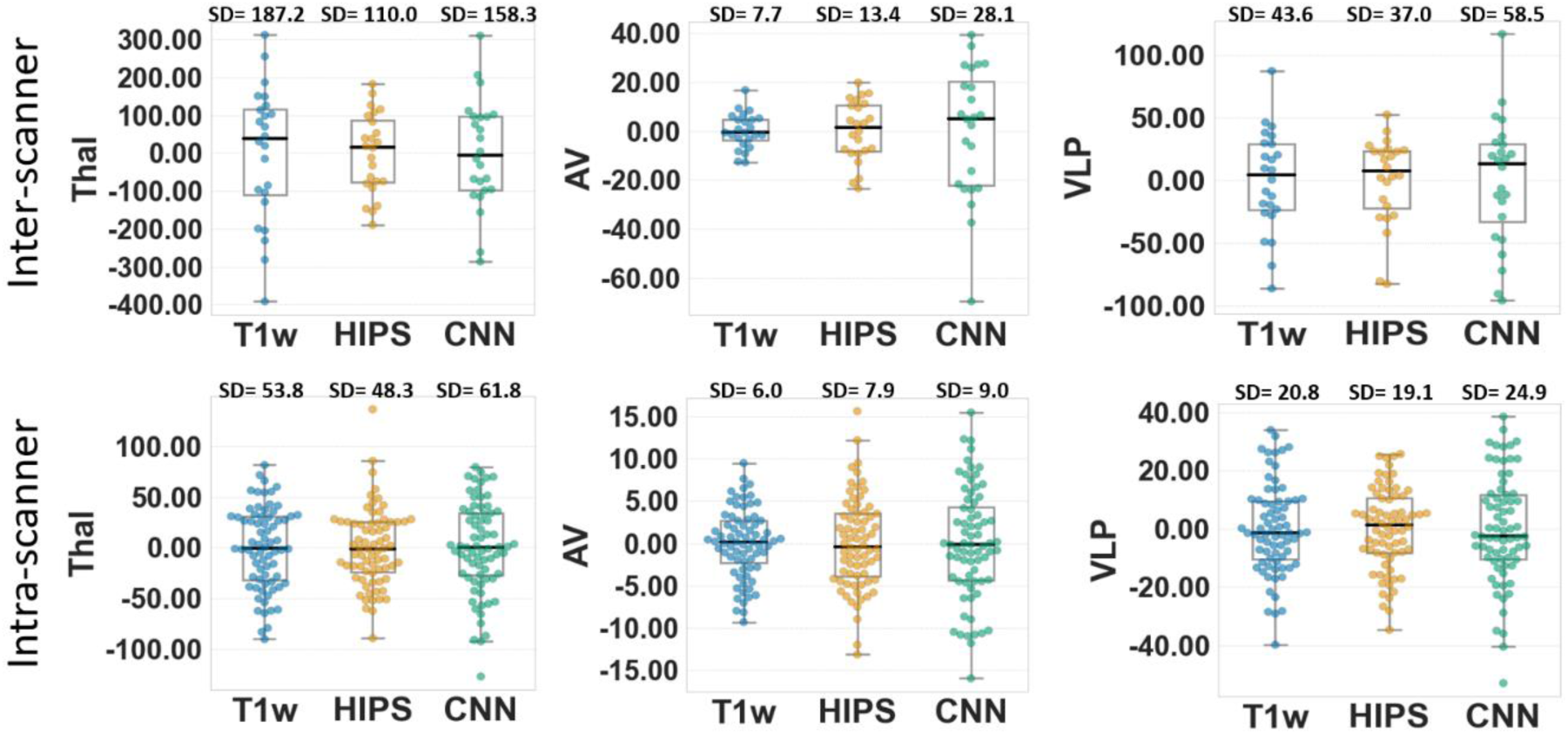
Inter-scanner and intra-scanner variability for the Frequently Traveling Human Phantom MRI dataset (in units of mm3) for T1w-THOMAS, HIPS-THOMAS, and CNN segmentation of the whole thalamus (Thal), AV and VLp nuclei. For this analysis, 24 different scanners covering 3 manufacturers, 2 field strengths and 4 sites each with 3 repeat scans per site were used, resulting in a dataset of 72 scans. Standard deviations (SD) for each method is shown above each plot.

## 4. Discussion

We have significantly improved the THOMAS pipeline which was developed for 7T WMn-MPRAGE data to allow thalamic nuclei segmentation from standard 3T T1w images, by employing a computationally efficient polynomial synthesis transform to generate WMn-like images prior to segmentation. Robustness of segmentations using HIPS-THOMAS and a single cubic function estimated from aggregating 10 3T datasets was demonstrated from data across differing T1 image contrasts (i.e. SPGR, MPRAGE, MP2RAGE), scanner manufacturers (i.e. Siemens, GE, Philips), and field strengths (i.e. 3T, 7T). HIPS-THOMAS had higher Dice coefficients, lower volume errors, and lower inter-scanner variability compared to T1w-THOMAS and CNN for most of the nuclei.

Segmentations of T1w data using HIPS-THOMAS are closer than T1w-THOMAS to THOMAS segmentations using WMn-MPRAGE images, resulting in improved volume accuracy and Dice coefficients for most of the nuclei. Dice was most improved for ventral nuclei (VA, VLp, VPL) where the improved contrast in WMn-like images better delineates external boundaries. The HIPS-THOMAS Dice improvement reaches its highest performance at 7T, likely due to the more accurate synthesis of WMn-like images at 7T using MP2RAGE T1 maps. Only the Hb nucleus has a decreased Dice index using HIPS-THOMAS on Siemens 7T data compared to the use of T1w-THOMAS (although not statistically significant), which may be explained by a higher variability of segmentations for this nucleus and a small effective in this dataset. These results are analogous to the results of Umapathy et al. (2021) where the synthesis-segmentation CNN displayed better accuracy compared to a segmentation network trained directly on T1w images, which showed spurious VPL atrophy. Results from the FTHP MRI dataset indicate that HIPS-THOMAS showed the least inter-scanner variability for the whole-thalamus and most nuclei. T1w-THOMAS, however, had lower intra-scanner variability for more nuclei than HIPS-THOMAS. This could be because the FTHP dataset included 1.5T MRI data which may not be optimal for HIPS or CNN. The standard deviation for intra-scanner variability for the whole thalamus using HIPS-THOMAS is comparable to the values reported in Opfer et al. (2023) using their custom CNN method (which only segments the whole thalamus and not nuclei) and FastSurfer (48 mm^3^ HIPS vs. 40 mm^3^ Opfer CNN, 45 mm^3^ FastSurfer). The standard deviation for inter-scanner variability for the whole thalamus using HIPS-THOMAS is significantly lower than the values reported in Opfer et al. (2023) (110 mm^3^ HIPS vs. 140 mm^3^ Opfer CNN, 310 mm^3^ FastSurfer).

This work used data from 3T Philips subjects to compute an aggregate 3^rd^ order function that was used for all subjects. The use of just Philips data is perhaps a limitation and linked to the non-availability of concurrent WMn-T1 data from other scanners at the commencement of this project. However, a comparison of this Philips-derived function to a function derived from multiple scanners found no significant differences in accuracy as discussed in Results (Supp. Tab. 3, Supp. Fig. 2). At 7T, the MP2RAGE sequence produces significantly higher contrast images than MPRAGE, resulting in synthesized WMn-like images that very closely match the WMn-MPRAGE images as seen by the near perfect linear density plot with minimal spread in Figure 2F. The 3^rd^ order polynomial performed optimally at 3T. However, at 7T, the linear function performed better, and this is reflected by the almost perfect unity line of the joint distribution plots in Figure 2F as well as better metrics and synthesized WMn-like images (Supp. Tab. 3 and Supp. Fig. 2). Given that 3T is the most common field strength used for neuroimaging and that the Dice improvements using the 3^rd^ order function was still very significant at 7T, we made the 3^rd^ order function as default in HIPS-THOMAS. (Note that HIPS-THOMAS can be run with a user-specified polynomial order and 7T data should be run with linear mode to get the best results). Future work will examine if use of functions optimized for each scanner and field strength is more beneficial as the exact nature of the curve depends on the parameters of the T1-MPRAGE sequence used, even though our normalization process helps reduce some of that variability. The significant contribution of HIPS to the THOMAS pipeline is to create images with similar contrast profiles as WMn-MPRAGE images, allowing the use of CC metric for nonlinear registration to the WMn template, which is more accurate than the MI metric used in T1w-THOMAS (Andronache et al., 2008). The joint fusion algorithm used in HIPS-THOMAS (cf. majority voting in T1w-THOMAS), also likely contributed to increased label accuracy (Bernstein et al., 2021; Pfefferbaum et al., 2023). HIPS is computationally efficient, does not add much complexity to the image analysis pipeline of THOMAS, and does not require separate training for the different scenarios of scanner manufacturers and field strengths, as required by CNN-based methods.

The CNN-synthesized images have less noise and slightly enhanced contrast compared to HIPS images (Fig. 3) due to denoising inherent in the synthesis CNN. In contrast, the use of polynomial functions in HIPS could result in noise amplification. Future work could address this using denoising methods. CNN performs comparably to HIPS-THOMAS on sequences on which it was trained (i.e. Siemens 3T MPRAGE) or on similar contrast images (SPGR), but HIPS-THOMAS is considerably better than the CNN for Philips 3T and Siemens 7T data, where the CNN failed on most cases due to lack of adequate training. While HIPS-THOMAS can yield higher Dice coefficients, it may paradoxically have higher volume errors than the CNN for certain nuclei on Siemens 3T (e.g. VPL) and GE 3T data (e.g. AV, LGN), as illustrated in Figure 8. This discrepancy may be due to the denoising present in CNNs, which allows for more precise delimitation. In summary, HIPS-THOMAS is more flexible and generalizable, as it can be applied easily to T1w images from different scanners without requiring training, a big advantage considering public databases like ADNI and OASIS contain data from a mix of manufacturers and field strengths.

Our work had some limitations. The T1w segmentations were evaluated against segmentations from THOMAS applied to WMn-MPRAGE images, which is not ideal. THOMAS has been thoroughly validated against manual segmentation at 3T (Su et al, ISMRM abstract) (Su et al., 2016) and 7T (Su et al., 2019) and was adopted as a “silver” standard, substituting for the “gold” standard manual segmentation, which is very time consuming and requires specific domain expertise. Another gold standard validation could be to use postmortem scanning in conjunction with histology to compare cytoarchitectonic features from histological staining to image contrasts from MRI. This however is very challenging due to many reasons including altered image contrasts due to the use of fixing agents post mortem and challenges of accurate registration between histology and MRI but would be very desirable for future validation work. The performance of the CNN method could also be enhanced by training the synthesis and segmentation networks with data from Philips and 7T scanners but was beyond the scope of this work. In the future, more sophisticated image and histogram normalization like that proposed in Nyul et al. (1999) or more complex exemplar-based synthesis approaches such as MIMECS (MR image example-based contrast synthesis) (Roy et al., 2011), could replace the simpler 3^rd^ order polynomial approach taken here, which could further improve HIPS performance and reduce noise amplification. This could also be useful for pipelines for analyzing whole brain images (as opposed to cropped images like in our current pipeline) where artifacts from scalp and other sources can impair HIPS performance. HIPS showed Dice improvements in several nuclei, but the Hb and VLa nuclei consistently showed < 0.7 Dice across all 3T scanners. While the poor habenula Dice could be attributed to its small size, the reasons for suboptimal VLa Dice compared to other ventral nuclei like the VA warrants further investigation.

## 5. Conclusion

WMn-like images synthesized using the computationally efficient HIPS significantly improved the robustness as well as the accuracy of THOMAS compared to direct THOMAS or CNN-based methods for segmentation of T1w data.

## 6. Data availability statement

The datasets analyzed during the current study are available from the corresponding author on reasonable request.

## 7. Code availability

The code used in the current study for generating the polynomial fits as well as the HIPS transformation and the integrated HIPS-THOMAS pipeline is available at https://github.com/thalamicseg/hipsthomasdocker.

## 9. Acknowledgements

We wish to acknowledge Germain Arribarat for his support and his time during the beginning of this project, forming J.P.V to image processing. This work was supported by the University of Toulouse 3 Paul Sabatier via a PhD student mobility grant obtained by J.P.V and the Swiss National Science Foundation. M.S would like to acknowledge funding from the National Institute of Biomedical Imaging and Bioengineering (7R01EB032674).

## 10. Author contributions

J.P.V. and M.S. conceived and developed the presented method. J.P.V. performed the computations and analysis under the supervision of M.S. J.P.V. wrote the manuscript with support from L.D., E.J.B., and mainly with M.S.

L.D., E.J.B., P.P., J.P., M.B.C., N.M.Z. acquired and shared MRI data from their previous respective studies and helped with the study design.

## 11. Additional information

Competing interests’ statement: The authors have no conflicts of interest relevant to this manuscript to disclose.

## 12. Supplementary information

**Supplementary Figure 1:**
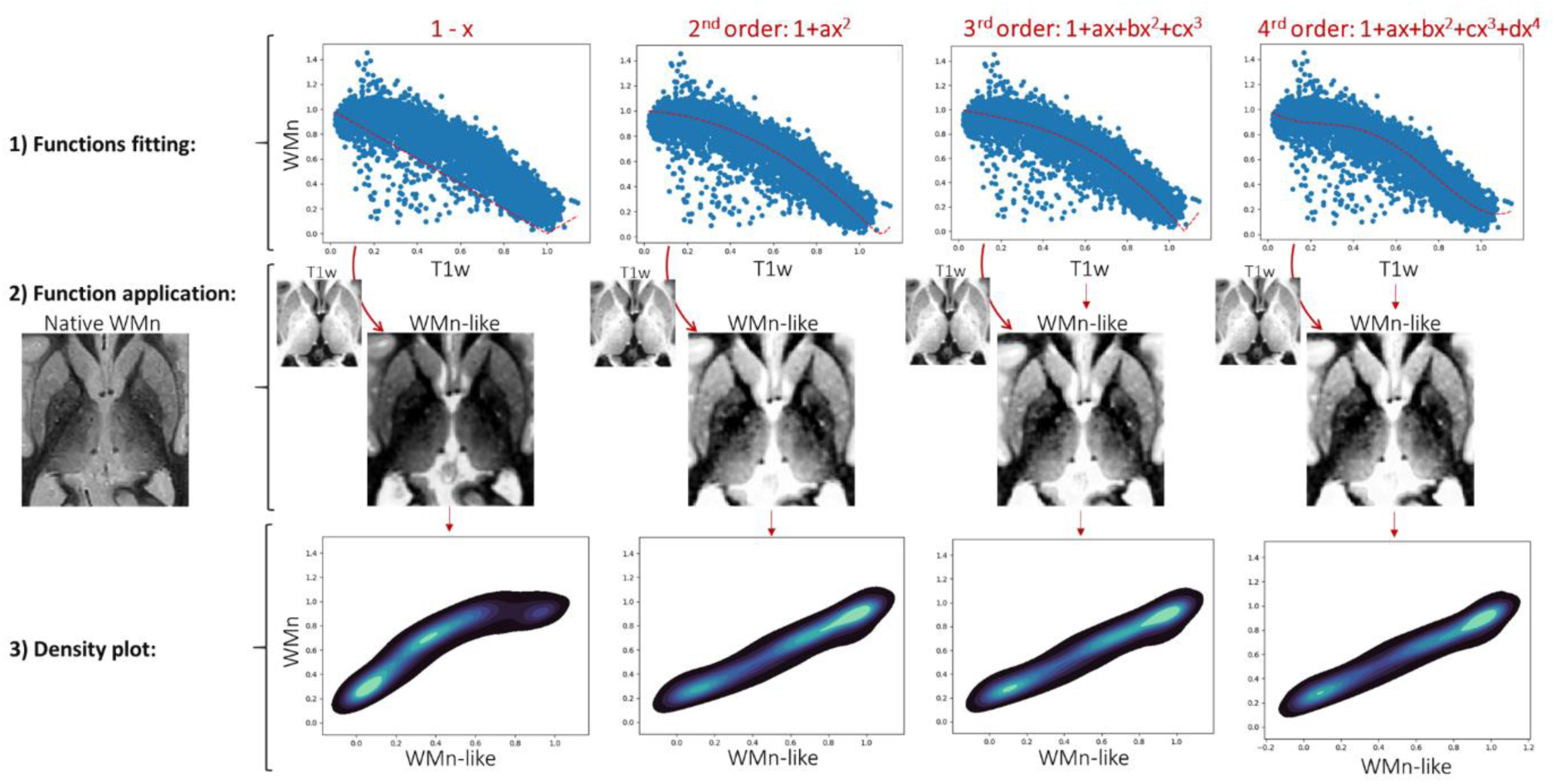
Plot of normalized T1w intensity vs. normalized WMn intensity along with polynomial fits overlaid (in red) for orders 1, 2, 3, and 4 (top row) for an example case. The higher order polynomials fit the data much better than the simple linear 1^st^ order. The corresponding transformed image (labeled WMn-like) is shown in the middle row for each order. Note the dark appearance of the transformed image for order 1. The density plots are shown in the bottom row comparing true WMn with WMn-like images generated by polynomial transformation of T1w.

**Supplementary Table 1:** Details of sequence parameters for MRI datasets used. *Images were synthesized using the inversion recovery equation (1-2 exp(-TI/T1) with TI=670ms and T1w images are generated using the T1 maps produced from MP2RAGE acquisition.

|  |  | Philips | GE | SIEMENS (1) | SIEMENS (2) |
| --- | --- | --- | --- | --- | --- |
| *Data source* |  | [1] | [2] | [3] | [4] |
| *N* |  | 18 | 19 | 12 | 9 |
| *Field strength* |  | 3T | 3T | 3T | 7T |
| *Sequence* |  | MPRAGE | SPGR | MPRAGE | MP2RAGE* |
| *T1w* | Voxel size (mm) | 0.9x0.9x1 | 0.9x0.9x1 | 1x1x1 | 0.8x0.8x0.8 or 0.6x0.6x0.6 |
|  | Flip angle (°) | 8 | 9 | 12 | 7 or 5 |
|  | TR/TE | 8.2/3.7 | 6.008/1.952 | 2000/2.52 | 6000/2.64 or 2.05 |
|  | Slice number | 170 | 120 | 192 | 176 or 320 |
|  | Matrix | 256x256 | 200x200 | 256x256 | 240x256 or 256x320 |
| *WMn* | Voxel size (mm) | 0.68x0.68x0.7 | 0.9x0.9x1.3 | 1x1x1 | * |
|  | Flip angle (°) | 8 | 7 | 7 |  |
|  | TR/TE | 9.7/4.7 | 11/5 | 4000/3.75 |  |
|  | Slice number | 120 | 210 | 160 |  |
|  | Matrix | 336x336 | 200x200 | 256x256 |  |

**Supplementary Table 2:** Mean SSI (structural similarity index) and MSE (mean square error) between the synthesized WMn image and the original WMn image of the same subject by applying a linear (1-x), 2^nd^ order, 3^rd^ order, and 4^th^ order function on its T1w image. This was applied on 10 Philips 3T MPRAGE, 9 GE 3T SPGR, 10 Siemens 3T MPRAGE and 8 Siemens 7T MP2RAGE cases. The aggregated function is computed by fitting the equation on 10 Philips cases, allowing more generalization on other data. The optimal functions for each order are listed in the HIPS parameter optimization section of Results.

|  |  | Linear | 2<sup>nd</sup> order | 3<sup>rd</sup> order | 4<sup>th</sup> order |
| --- | --- | --- | --- | --- | --- |
| PHILIPS 3T | SSI | 0.53 | **0.66** | **0.66** | 0.64 |
|  | MSE | 0.06 | 0.02 | 0.02 | 0.02 |
| GE 3T | SSI | 0.53 | **0.71** | **0.71** | 0.65 |
|  | MSE | 0.08 | 0.02 | 0.02 | 0.03 |
| SIEMENS 3T | SSI | 0.48 | **0.63** | **0.63** | 0.57 |
|  | MSE | 0.09 | 0.04 | 0.04 | 0.05 |
| SIEMENS 7T | SSI | **0.79** | 0.72 | 0.71 | 0.69 |
|  | MSE | 0.02 | 0.04 | 0.04 | 0.04 |

**Supplementary Figure 2:**
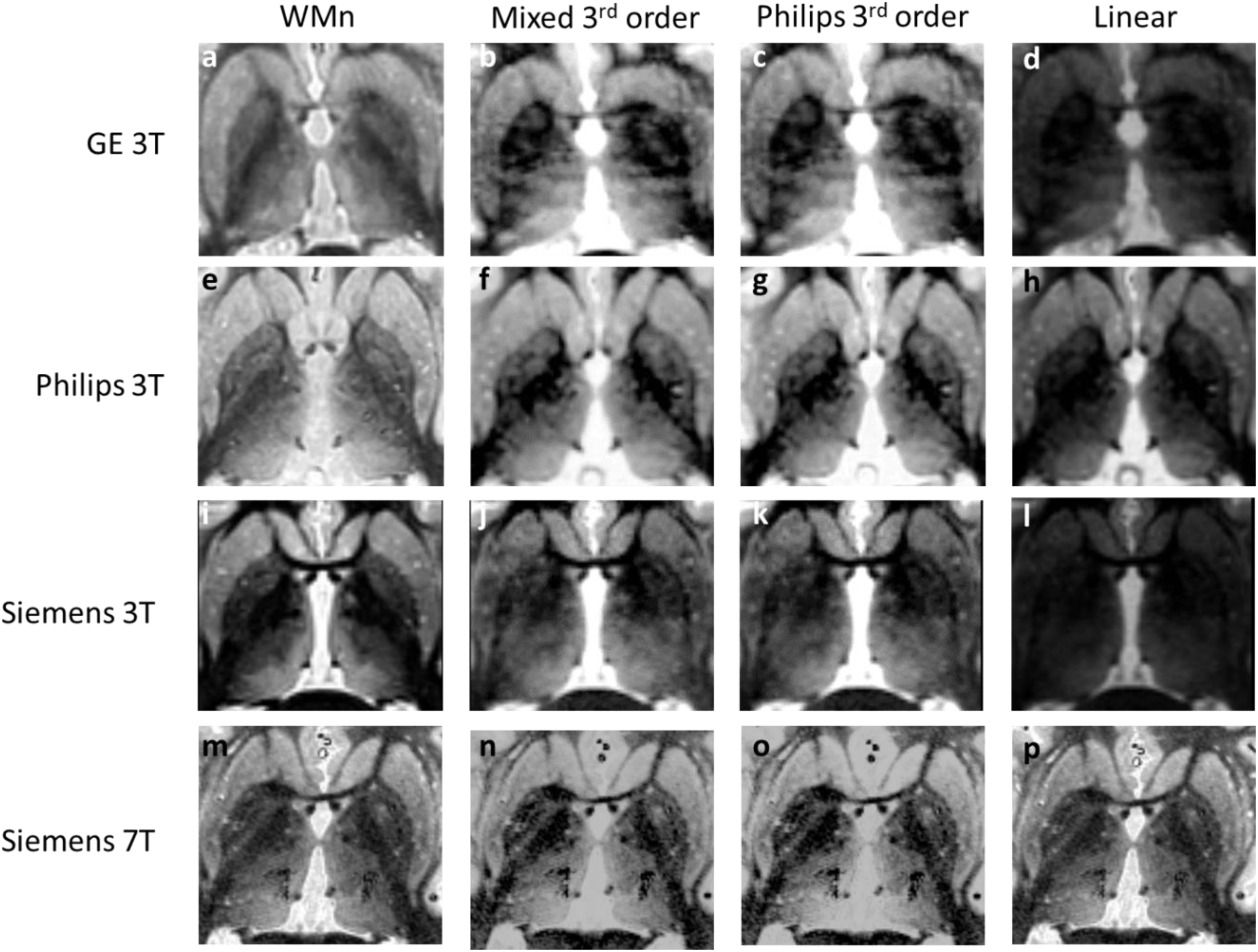
An axial slice from native WMn-MPRAGE compared with WMn-like images resulting from the application of the Philips averaged function, mixed dataset function (see Supplementary Table 7 for details), or linear (1-x) function on an example GE 3T SPGR (a-d), Philips 3T MPRAGE (e-h), Siemens 3T MPRAGE (i-l) or Siemens 7T MP2RAGE (m-p) subject. Note the similarity of the transformation when applying the mixed (b, f, j, n) and Philips (c, g, k, o) function on native T1w-images. Note also the optimality of the linear function for Siemens 7T data (p).

**Supplementary Table 3:** Mean Structural Similarity Index (SSI) and Mean Square Error (MSE) measures when comparing an axial slice native WMn and synthesized WMn images after applying a quadratic and cubic functions estimated from a mixed dataset comprising of 10 Philips 3T MPRAGE, 9 GE 3T SPGR, 10 Siemens 3T MPRAGE and 8 Siemens 7T MP2RAGE cases. The aggregated function is computed by fitting the equation on mixed data. For comparison, the SSI and MSE from using just 2^nd^ and 3^rd^ order Philips 3T cases is shown. The two functions perform comparably.

|  |  | Mixed<br>2<sup>nd</sup> order | Mixed<br>3<sup>rd</sup> order | Philips<br>2<sup>nd</sup> order | Philips<br>3<sup>rd</sup> order |
| --- | --- | --- | --- | --- | --- |
| PHILIPS 3T | SSI | 0.66 | 0.67 | 0.66 | 0.66 |
|  | MSE | 0.02 | 0.04 | 0.02 | 0.02 |
| GE 3T | SSI | 0.69 | 0.70 | 0.71 | 0.71 |
|  | MSE | 0.02 | 0.02 | 0.02 | 0.02 |
| SIEMENS 3T | SSI | 0.61 | 0.61 | 0.63 | 0.63 |
|  | MSE | 0.03 | 0.03 | 0.04 | 0.04 |
| SIEMENS 7T | SSI | 0.75 | 0.77 | 0.72 | 0.71 |
|  | MSE | 0.03 | 0.03 | 0.04 | 0.04 |

**Supplementary Table 4:**
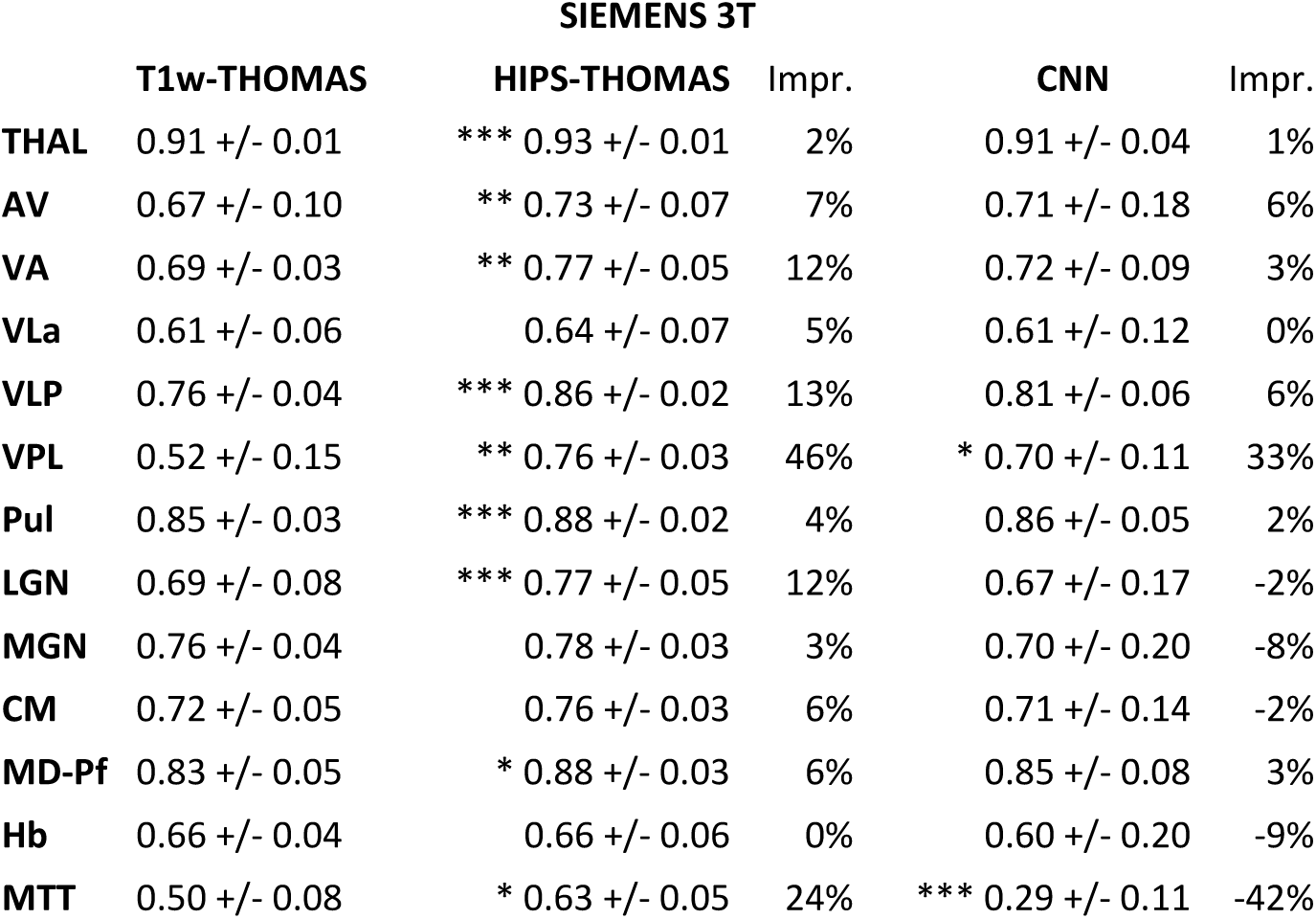
Mean Dice +/- SD of T1w-THOMAS, HIPS-THOMAS, and CNN segmentations along with % improvement for HIPS-THOMAS and CNN compared to T1w-THOMAS for 3T Siemens MPRAGE (n=12) data. HIPS-THOMAS improves Dice by >10% in 4 nuclei and MTT compared to 1 nucleus for CNN. P-values *<0.00385 (Bonferonni correction for multiple comparisons, 0.05/13) **<0.001 ***<0.0001.

**Supplementary Table 5:** Mean Dice +/- SD of T1w-THOMAS, HIPS-THOMAS, and CNN segmentations along with % improvement for HIPS-THOMAS and CNN compared to T1w-THOMAS for GE 3T SPGR data (n=19). HIPS-THOMAS improves Dice by >15% in 5 nuclei and MTT compared to 1 nucleus for CNN. P-values *<0.00385 (Bonferonni correction for multiple comparisons, 0.05/13) **<0.001 ***<0.0001.

| GE 3T |  |  |  |  |  |
| --- | --- | --- | --- | --- | --- |
|  | T1w-THOMAS | HIPS-THOMAS | Impr. | CNN | Impr. |
| **THAL** | 0.91 +/- 0.01 | *** 0.93 +/- 0.01 | 2% | 0.92 +/- 0.01 | 1% |
| **AV** | 0.68 +/- 0.09 | 0.69 +/- 0.12 | 2% | 0.70 +/- 0.08 | 4% |
| **VA** | 0.70 +/- 0.06 | *** 0.81 +/- 0.02 | 15% | * 0.76 +/- 0.03 | 12% |
| **VLa** | 0.64 +/- 0.08 | 0.66 +/- 0.06 | 4% | 0.62 +/- 0.08 | -2% |
| **VLP** | 0.74 +/- 0.08 | *** 0.85 +/- 0.03 | 16% | *** 0.81 +/- 0.04 | 12% |
| **VPL** | 0.56 +/- 0.14 | *** 0.78 +/- 0.06 | 40% | *** 0.72 +/- 0.07 | 40% |
| **Pul** | 0.85 +/- 0.03 | *** 0.89 +/- 0.02 | 5% | 0.87 +/- 0.02 | 2% |
| **LGN** | 0.67 +/- 0.04 | *** 0.78 +/- 0.04 | 17% | *** 0.75 +/- 0.04 | 13% |
| **MGN** | 0.78 +/- 0.04 | 0.82 +/- 0.04 | 5% | 0.69 +/- 0.10 | -9% |
| **CM** | 0.67 +/- 0.09 | *** 0.77 +/- 0.06 | 15% | * 0.75 +/- 0.03 | 11% |
| **MD-Pf** | 0.81 +/- 0.05 | *** 0.88 +/- 0.02 | 8% | * 0.86 +/- 0.02 | 6% |
| **Hb** | 0.66 +/- 0.07 | 0.70 +/- 0.04 | 6% | 0.67 +/- 0.12 | 3% |
| **MTT** | 0.47 +/- 0.10 | *** 0.63 +/- 0.05 | 32% | ** 0.31 +/- 0.08 | -34% |

**Supplementary Table 6:**
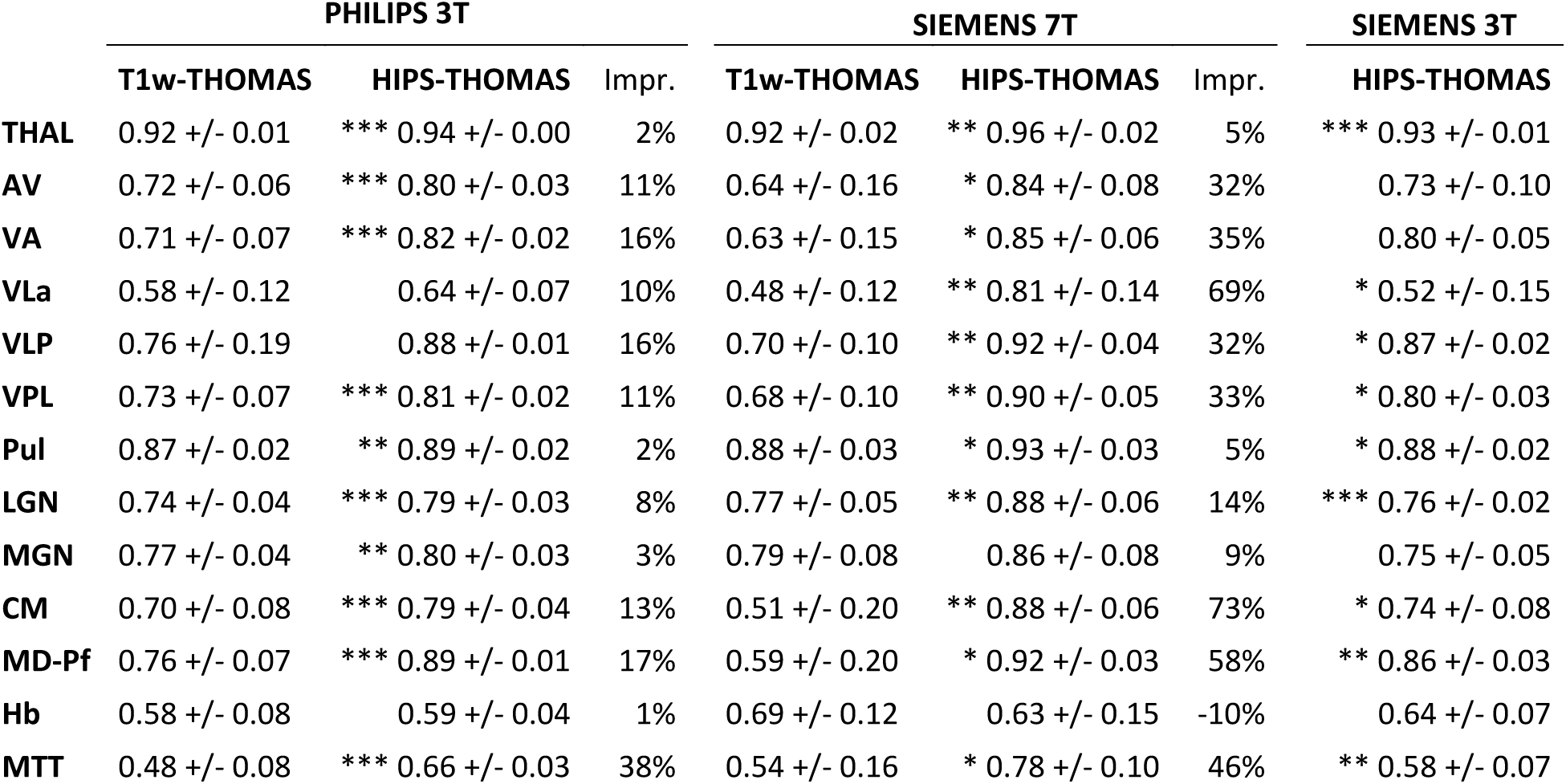
Mean Dice coefficient +/- SD of T1w-THOMAS and HIPS-THOMAS segmentations along with % improvement for HIPS-THOMAS compared to T1w-THOMAS for Philips 3T MPRAGE (n=18) and Siemens 7T MP2RAGE (n=8) data. HIPS-THOMAS improves Dice by >10% in 6 nuclei and MTT on Philips 3T data and by >30% in 7 nuclei and MTT on Siemens 7T data. The rightmost column shows Dice results for Siemens 3T MPRAGE data obtained on the same 8 subjects scanned on Siemens 7T. P-values *<0.00385 (Bonferonni correction for multiple comparisons, 0.05/13) **<0.001 ***<0.0001.

|  | PHILIPS 3T |  |  | SIEMENS 7T |  |  | SIEMENS 3T |
| --- | --- | --- | --- | --- | --- | --- | --- |
|  | T1w-THOMAS | HIPS-THOMAS | Impr. | T1w-THOMAS | HIPS-THOMAS | Impr. | HIPS-THOMAS |
| **THAL** | 0.92 +/- 0.01 | *** 0.94 +/- 0.00 | 2% | 0.92 +/- 0.02 | ** 0.96 +/- 0.02 | 5% | *** 0.93 +/- 0.01 |
| **AV** | 0.72 +/- 0.06 | *** 0.80 +/- 0.03 | 11% | 0.64 +/- 0.16 | * 0.84 +/- 0.08 | 32% | 0.73 +/- 0.10 |
| **VA** | 0.71 +/- 0.07 | *** 0.82 +/- 0.02 | 16% | 0.63 +/- 0.15 | * 0.85 +/- 0.06 | 35% | 0.80 +/- 0.05 |
| **VL<sub>a</sub>** | 0.58 +/- 0.12 | 0.64 +/- 0.07 | 10% | 0.48 +/- 0.12 | ** 0.81 +/- 0.14 | 69% | * 0.52 +/- 0.15 |
| **VLP** | 0.76 +/- 0.19 | 0.88 +/- 0.01 | 16% | 0.70 +/- 0.10 | ** 0.92 +/- 0.04 | 32% | * 0.87 +/- 0.02 |
| **VPL** | 0.73 +/- 0.07 | *** 0.81 +/- 0.02 | 11% | 0.68 +/- 0.10 | ** 0.90 +/- 0.05 | 33% | * 0.80 +/- 0.03 |
| **Pul** | 0.87 +/- 0.02 | ** 0.89 +/- 0.02 | 2% | 0.88 +/- 0.03 | * 0.93 +/- 0.03 | 5% | * 0.88 +/- 0.02 |
| **LGN** | 0.74 +/- 0.04 | *** 0.79 +/- 0.03 | 8% | 0.77 +/- 0.05 | ** 0.88 +/- 0.06 | 14% | *** 0.76 +/- 0.02 |
| **MGN** | 0.77 +/- 0.04 | ** 0.80 +/- 0.03 | 3% | 0.79 +/- 0.08 | 0.86 +/- 0.08 | 9% | 0.75 +/- 0.05 |
| **CM** | 0.70 +/- 0.08 | *** 0.79 +/- 0.04 | 13% | 0.51 +/- 0.20 | ** 0.88 +/- 0.06 | 73% | * 0.74 +/- 0.08 |
| **MD-Pf** | 0.76 +/- 0.07 | *** 0.89 +/- 0.01 | 17% | 0.59 +/- 0.20 | * 0.92 +/- 0.03 | 58% | ** 0.86 +/- 0.03 |
| **Hb** | 0.58 +/- 0.08 | 0.59 +/- 0.04 | 1% | 0.69 +/- 0.12 | 0.63 +/- 0.15 | -10% | 0.64 +/- 0.07 |
| **MTT** | 0.48 +/- 0.08 | *** 0.66 +/- 0.03 | 38% | 0.54 +/- 0.16 | * 0.78 +/- 0.10 | 46% | ** 0.58 +/- 0.07 |

**Supplementary Table 7:** Intra-scanner and inter-scanner variability methods for the Frequently Traveling Human Phantom MRI dataset [5] reported as standard deviation (in units of mm3) for T1w-THOMAS, HIPS-THOMAS, and CNN segmentation. For this analysis, 24 different scanners covering 3 manufacturers, 2 field strengths and 4 sites each with 3 repeat scans per site were used, resulting in a dataset comprised of 72 scans.

|  |  | *Thal* | *AV* | *VA* | *Vla* | *VLp* | *VPL* | *Pul* | *LGN* | *MGN* | *CM* | *MD* | *Hb* | *MTT* |
| --- | --- | --- | --- | --- | --- | --- | --- | --- | --- | --- | --- | --- | --- | --- |
| *INTRA*<br>(mm<sup>3</sup>) | T1w | 53.8 | **6.0** | **8.8** | 5.6 | 20.8 | **12.2** | **25.4** | **3.7** | **3.2** | **5.2** | **16.0** | **2.1** | 2.8 |
|  | HIPS | **48.3** | 7.9 | **8.8** | **4.2** | **19.1** | 13.4 | 29.7 | 6.8 | **3.1** | 9.0 | 24.2 | 2.6 | 2.9 |
|  | CNN | 61.8 | 9.0 | 10.6 | 5.7 | 24.9 | 12.5 | 31.4 | 5.7 | 4.0 | 8.5 | 17.9 | 2.5 | **2.7** |
| *INTER*<br>(mm<sup>3</sup>) | T1w | 187.2 | **7.7** | 18.0 | 8.2 | 43.6 | 26.5 | 84.1 | **6.6** | 6.2 | **8.9** | 48.6 | **3.2** | **3.3** |
|  | HIPS | **110.0** | 13.4 | **13.4** | **4.6** | **37.0** | **20.7** | **62.6** | 11.0 | **4.3** | **9.1** | **30.7** | 3.7 | 4.3 |
|  | CNN | 158.3 | 28.1 | 20.8 | 13.0 | 58.5 | 28.7 | 96.9 | 13.5 | 7.8 | 15.6 | 51.4 | 4.1 | 5.1 |

